# Returning Genomic Dietary Data to Participants by Translating Stool DNA into a Personal Food Profile

**DOI:** 10.64898/2026.07.29.26359128

**Authors:** Kylie R. Jeffrey, Caroline N. Rivera, Ammara Aqeel, Matthew J. Gedye, Nolan W. Ives, Sharon Jiang, Michelle C. Kirtley, Anna E. Bauer, Lawrence A. David

**Author notes:** Correspondence: Kylie R. Jeffrey and Lawrence A. David. denotes equal contribution (Kylie R. Jeffrey, Caroline N. Rivera). denotes co-corresponding author.

## Abstract

Returning individual results to research participants is increasingly expected in genomic studies. Yet, genomic dietary data have unique characteristics. New DNA sequencing techniques reconstruct diet from the residual plant and animal DNA recoverable in stool, reporting not nutrients or calories but a list of the species a person ate. Turning that list into something a participant can understand does not yet have an established framework. The Everyone EATS (Edible Atlas Through Sequencing) study returned personalized genomic dietary profiles to a non-clinical pilot cohort. Participants collected a stool sample at home, samples were processed with FoodSeq, and each participant received an interactive Diet Data Return Report that translated detected taxa into familiar food groups and benchmarked them against prior FoodSeq cohorts. No monetary compensation was offered, and the Diet Data Return Report was the sole incentive. Of 111 kits mailed, 80 were returned (72.1%) and sequenced (100% sequencing success for plant DNA; 98.7% for animal). Among report recipients with engagement data (n = 73), all accessed their report and 74% engaged interactively with its content. Among 16 feedback survey completers, 13/16 (81.2%) found the report easy to understand, 13/16 (81.2%) reported improved understanding of their dietary variety, and 5/16 (31.2%) reported a dietary behavior change. Curiosity about personal dietary data and supporting broader research goals were the most common reasons for enrolling (79.8%, respectively). Notably, the foods participants flagged as missing or unexpected clustered among herbs, spices, seafood, and processed items, the foods most easily forgotten in self-report and least readily assigned from sequence. Overall, our results suggest that genomic dietary data can be returned in a form participants find understandable and engaging, and that curiosity alone can motivate participation without payment.

## 1. Introduction

The return of individual research results to study participants has become a defining challenge for genomic and precision health research (1, 2). Genomic dietary assessment represents a particularly underexplored case. DNA metabarcoding of stool can now reconstruct individual dietary patterns from biological specimens, generating objective, participant-independent dietary profiles without relying on self-report (3–5). Whether and how to return these data to participants, in a form they find understandable and worth engaging with, has not previously been examined.

A growing policy consensus has positioned the return of individual research results as both an ethical obligation and a strategy for sustaining participant engagement. The National Academies of Sciences, Engineering, and Medicine identified returning value to participants as a defining feature of a contemporary research paradigm in 2018 (2), and NIH has since called for and codified these expectations across funded programs (6, 7). Most of this guidance, however, has been developed around clinical genetic findings. Genomic dietary data are different in kind, and the specific challenge of communicating them has barely been addressed. This gap has become newly relevant with the emergence of FoodSeq (8) and MEDI (9), methods that can now reconstruct individual dietary intake from food-derived DNA in stool, independent of what participants remember eating.

FoodSeq reconstructs dietary intake by applying DNA metabarcoding to the residual plant and animal DNA recoverable from stool (3, 10). The method uses two complementary markers: the chloroplast *trnL*-P6 region captures plant-derived dietary DNA and yields plant metabarcoding richness (pMR), the count of distinct plant taxa per sample, which correlates with traditional dietary diversity metrics across observational cohorts (ρ = 0.40 to 0.63) (3); the 12S mitochondrial rRNA V5 region captures vertebrate animal-derived DNA (11). Thus, from a stool sample, FoodSeq characterizes diet across all major food groups without relying on participant recall.

However, returning these data to participants presents a communication challenge with few direct precedents. The closest analogy comes from microbiome research, where stool-derived sequencing data have been returned to participants for years (12), but a food profile differs from a microbiome report in a key respect: it names foods, entities participants recognize and can act on (8, 13), rather than bacteria most people cannot place. Despite this interpretive advantage, a FoodSeq profile is not a conventional dietary report. It returns no calories, macronutrients, or serving sizes; instead, it yields a list of detected taxa, each of which may correspond to multiple edible species within a genus or family. Translating that output into something a participant can understand requires an interpretive layer for which no standard yet exists.

Here we describe the Everyone EATS (Edible Atlas Through Sequencing) study, in which we returned personalized genomic dietary profiles to participants in a fully remote pilot cohort. Although FoodSeq has been applied across several cohorts, the return of FoodSeq data to participants has not previously been reported. No monetary compensation was offered; access to a personalized dietary profile served as the sole incentive. We report initial evidence that genomic dietary data can be returned in a form participants find understandable and engaging, that curiosity about one’s own diet can motivate participation, and that the residual difficulty participants encountered points to translation from sequence to food, not sequencing, as the key unsolved step.

## 2. Methods

### 2.1 Study Design and Setting

Everyone EATS was a fully remote, citizen-science observational cohort study approved by the Duke University Health System Institutional Review Board (Pro00049498; PI: Dr. Lawrence David; study). No in-person visits were required; all study procedures were administered through REDCap (Research Electronic Data Capture). Recruitment was intentionally limited in scope during this pilot phase, consisting of physical flyers posted in five Duke University campus buildings, a single targeted email distributed to affiliates of the Duke University Forever Learning Institute (FLI) partner network, and internal university email lists; no active or social media recruitment was conducted. Participants received no monetary compensation; a personalized DDRR summarizing each participant’s genomic food profile served as the sole participant incentive.

### 2.2 Study Onboarding and Baseline Survey

Interested individuals completed a brief REDCap screening survey; the primary exclusion criterion was antibiotic use within the preceding 30 days, given its known disruption of gut DNA profiles (14). Eligible individuals completed electronic consent (eConsent) via REDCap, covering stool collection procedures, DDRR delivery, data privacy provisions, and withdrawal rights. Consented participants completed a REDCap baseline survey covering demographics, diet type, gastrointestinal history, supplement use, and lifestyle factors (**Supplementary Material**). All data were managed in REDCap and exported for analysis.

### 2.3 At-Home Sampling and FoodSeq Pipeline

#### 2.3.1 Sample Kits and Stool Collection

Participants who completed the baseline survey received a mail-based stool collection kit. Kit contents included stool collection supplies, cryo-safe labels, non-latex gloves, and a return box affixed with a prepaid shipping label addressed to the David Laboratory. Written collection and shipping instructions were supplemented by a linked instructional video (https://everyoneeatsproject.org/collection-kit). Participants were instructed to collect three scoops from a single bowel movement, label and seal the vials, and return the sample within 24 hours of collection; guidance was provided for cases requiring delayed shipping.

#### 2.3.2 FoodSeq Laboratory Processing

Received stool samples were processed using the FoodSeq pipeline as described previously (3, 11). Briefly, dietary DNA was extracted and amplified using two complementary markers: the chloroplast trnL-P6 region, which captures plant-derived dietary DNA, and the 12S mitochondrial rRNA V5 region, which captures vertebrate animal-derived dietary DNA. Libraries were sequenced on an Illumina MiniSeq. Reads were demultiplexed, quality-filtered, and mapped to a curated food reference database. The bioinformatic pipeline used to process FoodSeq data is publicly available at https://doi.org/10.5281/zenodo.20174235.

### 2.4 Personalized Data Return

#### 2.4.1 Overview of the Diet Data Return Report (DDRR)

Following successful sequencing and quality control, each participant received a personalized, interactive DDRR delivered via a unique hyperlink distributed through an automated REDCap email. We designed each component around what FoodSeq data can reliably support and around what participants may recognize from conventional nutrition reporting. Where possible, individual scores are situated within cohort benchmarks from prior FoodSeq studies, since a metric viewed alongside a distribution is more interpretable than one presented alone (15, 16). Each report included: (1) a list of food taxa detected in the participant’s stool sample, organized by food group; (2) a food group profile visualization comparing the participant’s genomic food profile to U.S. and international cohort benchmarks; (3) a food color diversity visualization illustrating detected plant taxa grouped by phytonutrient color category; (4) a dietary processing likelihood score; and (5) educational free-text sections contextualizing findings within population-level dietary science (**Figure 1; Supplementary Material**). Reports were generated using R and Python for data processing and the Infogram API for interactive visualization and were distributed in two batches (March 2026 and June 2026). Report generation was triggered manually by study staff.

**Figure 1.**
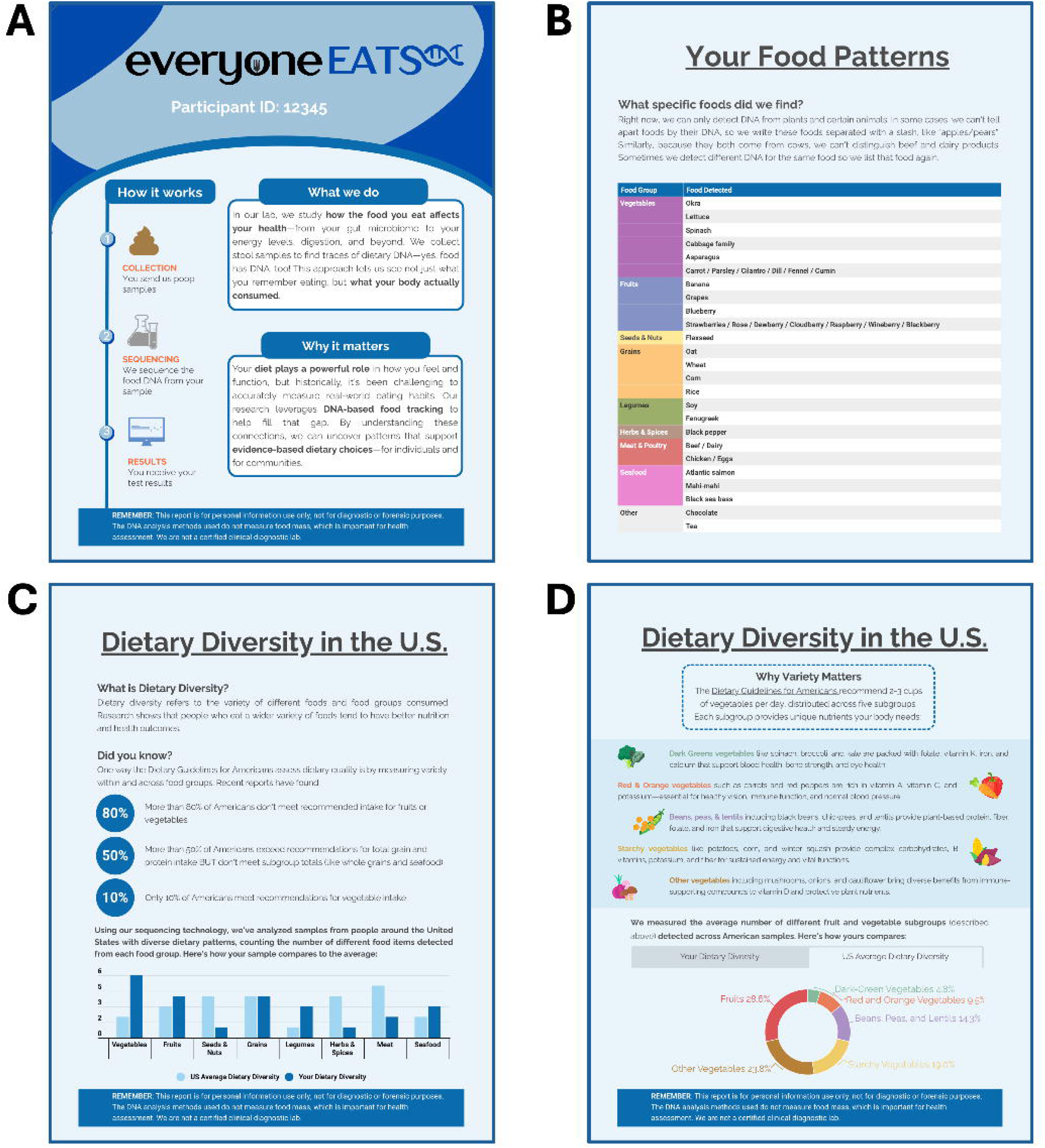
Example excerpt from a Diet Data Return Report. Representative panels from a personalized Diet Data Return Report (DDRR), illustrating a subset of the full report. (A) Introductory panel describing the FoodSeq workflow and its rationale. (B) Table of detected food taxa organized by food group, with taxonomic ambiguity indicated with using a slash-delimited format. (C-D) Dietary diversity comparison panels showing the participant’s food group counts relative to a U.S. cohort average, alongside contextual statistics on national dietary intake patterns from the 2020-2025 USDA Dietary Guidelines for Americans. Participant identifiers shown are illustrative and do not correspond to an actual study participant. The full DDRR included additional interactive elements not depicted here (**Supplementary Materials**).

#### 2.4.2 Translating Sequence into Food

Translating FoodSeq output into a participant-facing profile required mapping detected sequences to foods, the central interpretive step in returning these data. Raw FoodSeq output consists of a list of detected amplicon sequence variants (ASVs) with associated read counts, mapped to food taxa via a curated reference database (**Supplementary Methods**) (3). To make this output interpretable to participants, each detected taxon was assigned to one of eight food groups (Vegetables, Fruits, Grains, Legumes, Seeds and Nuts, Herbs and Spices, Meat and Poultry, and Seafood) using a curated lookup table. When an ASV resolved unambiguously to a single food type, the corresponding food group was assigned directly. When an ASV was attributable to multiple food types within a plant family (as occurs when the trnL-P6 marker does not distinguish among species within a genus or family), food group assignment was made by considering two factors: the food item most likely to be consumed in the U.S. population, and the food item most likely to be consumed in sufficient quantity to contribute detectable DNA to stool. The latter factor reflects a key property of metabarcoding data, namely that read presence is driven not only by dietary frequency but also by the mass of plant material ingested, which varies substantially across food types. Where these two factors pointed to the same food group, that group was assigned; where they diverged, a judgment among study team members was made weighing both considerations in context. For example, the family *Lauraceae* encompasses avocado (*Persea americana*), bay leaf (*Laurus nobilis*), and cinnamon (*Cinnamomum spp*.). Because avocado is consumed in substantially greater mass per eating occasion than bay leaf or cinnamon, *Lauraceae* detections were assigned to the Fruits group and listed as “Avocado / Bay Leaf / Cinnamon” in participants’ reports. This slash-delimited labeling convention was used in the participants’ personalized food taxa table to transparently communicate taxonomic ambiguity to participants while retaining a clear food group assignment.

Following food group assignment, a separate color mapping was applied to plant-derived taxa for the food color diversity visualization (17–19). Each plant taxon was assigned a color category based on its food group classification: Red (e.g., tomatoes, strawberries); Orange and Yellow (e.g., carrots, mangoes); Green (e.g., spinach, broccoli); Blue and Purple (e.g., blackberries, purple cabbage); and White and Brown (e.g., garlic, onions, mushrooms). The same two-factor logic applied to food group assignment (likelihood of consumption and likelihood of sufficient quantity for DNA detection) was used to resolve color assignments for taxa attributable to multiple plant types. This visualization was designed to connect FoodSeq output to color-based dietary diversity framework used in U.S. public health messaging. (18)

#### 2.4.3 Educational Free-Text Sections

The DDRR included educational free-text contextualizing FoodSeq technology and population-level dietary patterns. Content was developed by the study team drawing on publicly available guidance from the USDA Dietary Guidelines for Americans (18); sections covering dietary diversity recommendations and food group guidance were written to reflect this published guidance directly. Phytonutrient content descriptions associated with the food color diversity visualization were informed by general nutrition literature (20–23). All sections described population-level associations without making individualized dietary recommendations, consistent with the non-diagnostic nature of the DDRR, and included a standard disclaimer that the report is for personal information use only, is not for diagnostic or forensic purposes, and that DNA analysis does not measure food mass.

### 2.5 DDRR Engagement Tracking

Participant engagement with each DDRR was tracked using Infogram’s built-in analytics, from which we extracted total view count per report and interaction rate, defined as the percentage of viewers who engaged with interactive report elements including tooltip hovers, chart clicks, map navigation, and tab selections.

### 2.6 Feedback survey

All participants who received a DDRR were invited to complete a brief post-report REDCap feedback survey (**Supplementary Material**). Survey domains included: report comprehension; accuracy perception; behavioral impact; future participation intent; recommendation likelihood (5-point Likert scale); and willingness to pay for similar services. Open-text fields allowed participants to describe comprehension barriers, surprising food detections, and the motivational impact of receiving their results.

### 2.7 Statistical Analysis

#### 2.7.1 Study Endpoints

The pre-specified primary endpoints of this study were kit return rate and sequencing success rate, as measures of operational feasibility in a remote, uncompensated cohort. Pre-specified secondary endpoints included DDRR initial access rate, report comprehension (proportion rating the report easy to understand), and interactive engagement rate. These endpoints were not modified during the course of the study. Analyses of behavioral outcomes, participant motivations, and feedback survey responses were not pre-specified and are considered exploratory.

#### 2.7.2 Completion Rate and Study Population Analysis

We report N and percentage at each stage of the enrollment funnel from eligibility form initiation through feedback survey completion. Continuous participant characteristics are summarized as mean ± standard deviation (SD); categorical variables are reported as N (%). Among participants who received a collection kit, we compared baseline demographic characteristics between kit returners and non-returners using chi-square or Fisher’s exact test for categorical variables and independent-samples t-test or Mann-Whitney U test for continuous variables.

#### 2.7.3 FoodSeq Pipeline Performance

We summarize FoodSeq output to confirm that the pipeline produced coherent dietary data. We report the sequencing success rate (the proportion of returned samples meeting the minimum read-depth threshold) and the number of food taxa and food groups detected. As a face-validity check, we compared FoodSeq-detected animal food groups against self-reported diet type (omnivore, pescatarian, vegetarian, vegan). Formal validation of FoodSeq against reference intake measures was outside the scope of this study.

#### 2.7.4 Engagement and Feedback Survey Analysis

Given the small number of feedback survey respondents (n = 16), findings from the feedback survey instrument are exploratory; we report frequencies and proportions only and do not perform inferential statistical comparisons.

## 3. Results

### 3.1 Enrollment, Cohort, and Sample Return

Participants enrolled, completed home stool collection, and returned kits at high rates despite receiving no compensation. Of 153 individuals who initiated the eligibility screening form, 145 (94.8%) completed it, and 5 (3.4% of completers) were ineligible, all due to recent antibiotic use. Of 140 eligible individuals, 121 (86.4%) completed consent, and 111 of these had a complete mailing address and received a kit. Of the 111 kits mailed, 80 were returned, a 72.1% return rate, with a median return time of 13 days (IQR 8.0–19.5). All 80 returned samples were sequenced (100% sequencing success for plant DNA; 98.7% for animal). DDRRs were generated for 73 sequenced participants and delivered in batches (March and June 2026), a median of 149 days after kit receipt. The full funnel is shown in **Figure 2**.

**Figure 2.**
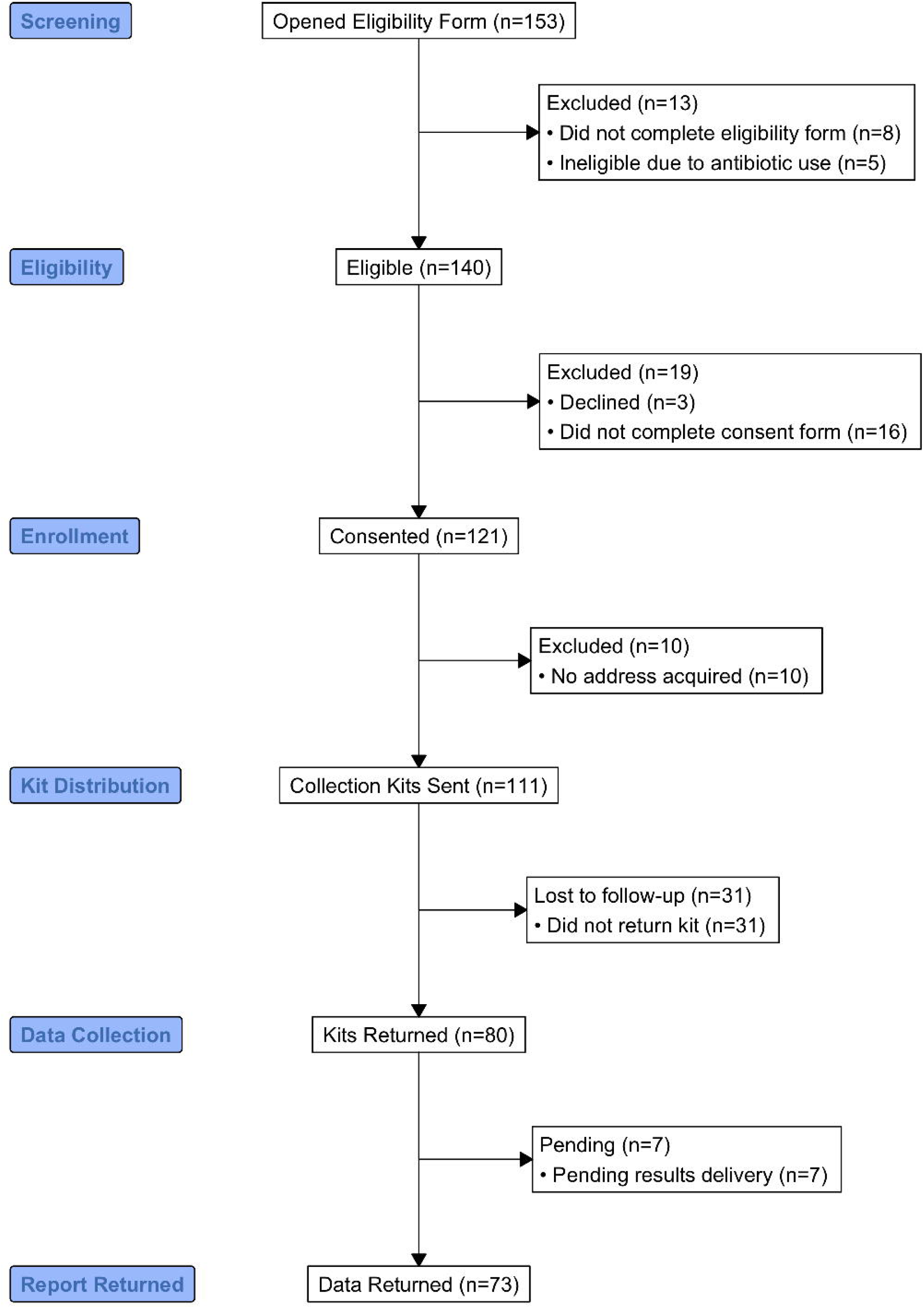
Study enrollment, kit return, and data return completion in the Everyone EATS pilot cohort. Flow diagram showing participant progression through the Everyone EATS study from initial screening through personalized data return. The diagram follows CONSORT reporting guidelines adapted for observational cohort studies.

The cohort showed signs of healthy volunteer bias (**Table 1**). Among the 109 adults who received a kit (2 minors were excluded from analysis), mean age was 44.2 ± 17.2 years (range 20–88); 78.0% were female and 85.3% White, 71.6% held a graduate or professional degree, and 55.9% reported annual household income at or above $100,000. Participants spanned 26 states but were concentrated near the study site (45.5% in North Carolina; **Figure 3**), and 13.8% were born outside the United States. Kit returners and non-returners differed on several characteristics (**Supplementary Table 1**). Returners were more likely to be female (87% vs. 63%, p = 0.005) and less likely to identify as Asian (9% vs. 27%, p = 0.027), and non-returners reported a higher prevalence of recent diarrhea (67% vs. 34%, p = 0.002) and trended younger (Mann-Whitney U p = 0.024; **Supplementary Table 1**).

**Figure 3.**
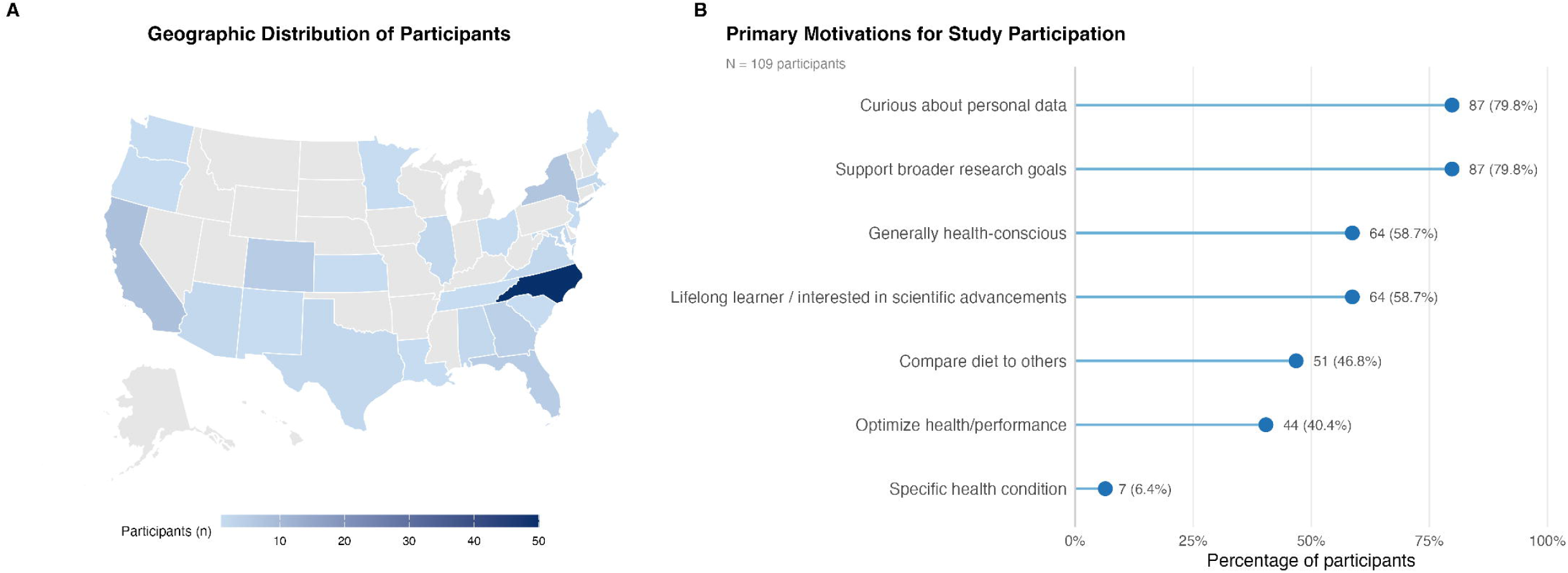
Geographic distribution and primary motivations for participation in the Everyone EATS study. (A) Geographic distribution of Everyone EATS participants. U.S. map showing participant counts by state (n=109 adults). Shading intensity represents number of participants per state. (B) Primary motivations for study participation. Horizontal bar chart showing percentage of participants endorsing each motivation category; multiple selections were permitted.

**Table 1.** Demographic and socioeconomic characteristics the Everyone EATS cohort. Characteristics of adult participants who received a stool collection kit (n=109 adults) and those who returned a kit (n=79 adults). Two participants under age 18 who received and returned a kit are excluded from all analysis.

| Variable | Kit Sent (n=109 adults) | Kit Returned (n=79 adults) |
| --- | --- | --- |
| **Age** |  |  |
| Mean ± SD (range) | 44.2 ± 17.2 (20-88) | 45.8 ± 16.1 (22-83) |
| **Sex Assigned at Birth** |  |  |
| Female | 88 (80.7%) | 69 (87.3%) |
| Male | 21 (19.3%) | 10 (12.7%) |
| **Gender Identity** |  |  |
| Female | 85 (78.0%) | 66 (83.5%) |
| Male | 20 (18.3%) | 10 (12.7%) |
| Non-binary/Other | 4 (3.7%) | 3 (3.8%) |
| **Race (multi-select)** |  |  |
| Asian | 15 (13.8%) | 7 (8.9%) |
| Black or African American | 2 (1.8%) | 1 (1.3%) |
| White | 93 (85.3%) | 70 (88.6%) |
| Other | 3 (2.8%) | 3 (3.8%) |
| **Hispanic or Latino** |  |  |
| Yes | 4 (3.7%) | 2 (2.5%) |
| No | 105 (96.3%) | 77 (97.5%) |
| **Country of Birth** |  |  |
| United States | 94 (86.2%) | 69 (87.3%) |
| Other country | 15 (13.8%) | 10 (12.7%) |
| **Education (highest level)** |  |  |
| Graduate or professional degree | 78 (71.6%) | 59 (74.7%) |
| Bachelor's degree | 21 (19.3%) | 14 (17.7%) |
| Some graduate or professional school | 8 (7.3%) | 5 (6.3%) |
| Associate's degree | 1 (0.9%) | -- |
| Some college or technical school | 1 (0.9%) | 1 (1.3%) |
| **Household Income** |  |  |
| \$150,000 and greater | 41 (37.6%) | 30 (38.0%) |
| \$100,000 to 149,999 | 20 (18.3%) | 15 (19.0%) |
| \$75,000 to 99,999 | 7 (6.4%) | 5 (6.3%) |

Returning Genomic Dietary Data to Participants by Translating Stool DNA into a Personal Food Profile Kylie R. Jeffrey and Caroline N. Rivera
| Variable | Kit Sent (n=109 adults) | Kit Returned (n=79 adults) |
| --- | --- | --- |
| \$50,000 to 74,999 | 9 (8.3%) | 6 (7.6%) |
| \$25,000 to 49,999 | 14 (12.8%) | 8 (10.1%) |
| Below \$25,000 | 2 (1.8%) | 1 (1.3%) |
| Prefer not to answer | 16 (14.7%) | 14 (17.7%) |
| **Living Situation (multi-select)** |  |  |
| Large city | 41 (37.6%) | 27 (34.2%) |
| Suburb near large city | 36 (33.0%) | 29 (36.7%) |
| Small city or town | 30 (27.5%) | 21 (26.6%) |
| Rural area | 4 (3.7%) | 3 (3.8%) |
| **Environment** |  |  |
| Tap water perceived as safe to drink | 97 (89.0%) | 72 (91.1%) |
| **Distance from Park** |  |  |
| 1 block | 26 (23.9%) | 16 (20.3%) |
| 2 blocks | 25 (22.9%) | 20 (25.3%) |
| ¼ mile | 18 (16.5%) | 14 (17.7%) |
| ½ mile or more | 39 (35.8%) | 28 (35.4%) |
| Prefer not to answer | 1 (0.9%) | 1 (1.3%) |
| **Food & Healthcare Access (past 12 months)** |  |  |
| Couldn't afford healthcare: Yes | 9 (8.3%) | 4 (5.1%) |
| Couldn't afford food: Yes | 1 (0.9%) | -- |

### 3.2 Participant Motivations

Enrollment was driven by curiosity rather than compensation. Curiosity about personal dietary data was the most commonly endorsed reason for joining (79.8%), tied with a desire to support broader research goals (79.8%). General health-consciousness (58.7%), intellectual interest in scientific advances (58.7%), interest in comparing one’s diet to others (46.8%), and optimizing health or performance (40.4%) were also frequently endorsed. The co-occurrence of personal curiosity and altruistic motivation indicates that participants were drawn both to learning about their own diet and to contributing to nutritional science (**Figure 3**).

### 3.3 Report Engagement

Recipients engaged with their reports repeatedly, not only on first access. All DDRR recipients accessed their report at least once (100% initial access). Among 73 recipients with available analytics, 71 (97.3%) viewed their report more than once (median 6 views per recipient), and 54 (74.0%) engaged interactively with report content beyond initial access (**Table 3**; **Figure 4**). This engagement occurred despite a median 149-day interval between sample collection and report delivery.

**Figure 4.**
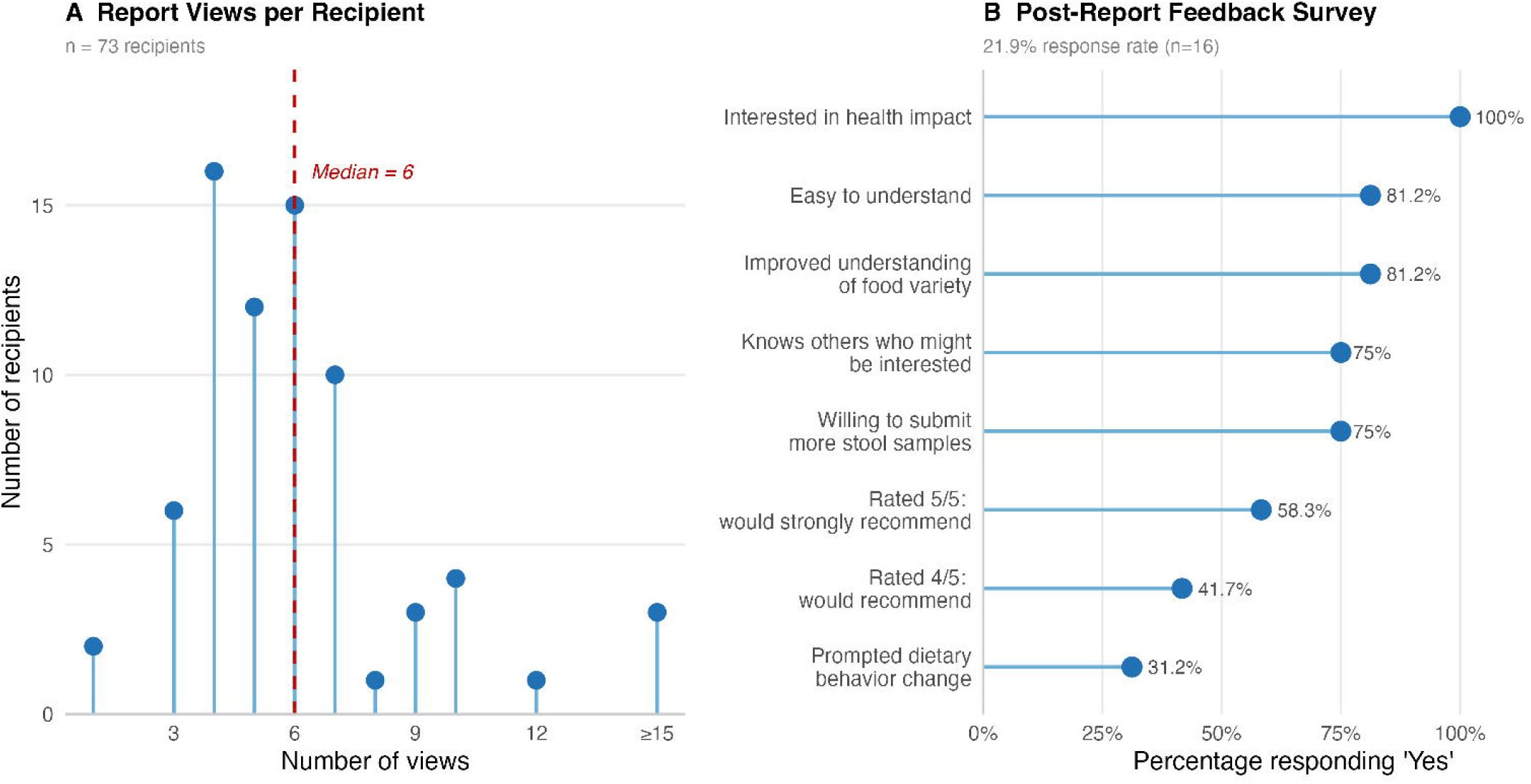
Participant engagement with Diet Data Return Reports and post-report feedback survey responses. (A) Report views per recipient. Distribution of the number of times each participant accessed their personalized Diet Data Return Report (DDRR) among recipients with available engagement data (n=73). Red dashed line indicates the median. (B) Post-report feedback survey responses. Horizontal bar chart showing percentage of feedback survey respondents (n=16) endorsing each survey item.

**Table 2.** Diet, health, and lifestyle characteristics of the Everyone EATS cohort. Dietary patterns, food purchasing behaviors, health history, and gastrointestinal characteristics of adult participants who received a stool collection kit (n = 109) and whose who returned a kit (n=79).

| Variable | Kit Sent (n=109 adults) | Kit Returned (n=79 adults) |
| --- | --- | --- |
| **Diet Type** |  |  |
| I eat anything with no exclusions (omnivore) | 90 (82.6%) | 63 (79.7%) |
| Vegetarian but eat fish | 10 (9.2%) | 8 (10.1%) |
| I eat anything except red meat | 3 (2.8%) | 3 (3.8%) |

Returning Genomic Dietary Data to Participants by Translating Stool DNA into a Personal Food Profile Kylie R. Jeffrey and Caroline N. Rivera
| Variable | Kit Sent (n=109 adults) | Kit Returned (n=79 adults) |
| --- | --- | --- |
| Vegetarian | 3 (2.8%) | 3 (3.8%) |
| Vegan | 3 (2.8%) | 2 (2.5%) |
| **Specialized Diets (multi-select)** |  |  |
| FODMAP | 1 (0.9%) | -- |
| Other low-grain/low-processed | 4 (3.7%) | 3 (3.8%) |
| Mediterranean | 5 (4.6%) | 5 (6.3%) |
| Exclude dairy | 7 (6.4%) | 6 (7.6%) |
| Exclude gluten | 6 (5.5%) | 5 (6.3%) |
| Exclude refined sugars | 3 (2.8%) | 3 (3.8%) |
| No specialized diet | 83 (76.1%) | 58 (73.4%) |
| Exclude nightshades | 1 (0.9%) | 1 (1.3%) |
| **Regular Beverage Consumption (any frequency)** |  |  |
| Coffee | 89 (81.7%) | 65 (82.3%) |
| Alcohol | 66 (60.6%) | 49 (62.0%) |
| **Food Purchasing Locations (past 30 days, multi-select)** |  |  |
| Trader Joe's | 59 (54.1%) | 42 (53.2%) |
| Whole Foods | 46 (42.2%) | 36 (45.6%) |
| Harris Teeter | 43 (39.4%) | 30 (38.0%) |
| Wholesale clubs (Costco/Sam's) | 37 (33.9%) | 25 (31.6%) |
| Target | 27 (24.8%) | 23 (29.1%) |
| Local farmers market | 37 (33.9%) | 28 (35.4%) |
| Aldi | 23 (21.1%) | 18 (22.8%) |
| Walmart | 29 (26.6%) | 21 (26.6%) |
| Local international grocery | 22 (20.2%) | 12 (15.2%) |
| Online grocery services | 14 (12.8%) | 8 (10.1%) |
| Food Lion | 21 (19.3%) | 16 (20.3%) |
| Co-op | 20 (18.3%) | 15 (19.0%) |
| Kroger | 14 (12.8%) | 13 (16.5%) |
| Wegmans | 21 (19.3%) | 18 (22.8%) |
| Publix | 19 (17.4%) | 15 (19.0%) |
| Fresh Market | 11 (10.1%) | 10 (12.7%) |
| Gas stations | 9 (8.3%) | 6 (7.6%) |
| H-mart | 6 (5.5%) | 3 (3.8%) |
| Dollar stores | 4 (3.7%) | 3 (3.8%) |
| Drug stores (CVS/Walgreens) | 3 (2.8%) | 2 (2.5%) |
| Lidl | 10 (9.2%) | 6 (7.6%) |

Returning Genomic Dietary Data to Participants by Translating Stool DNA into a Personal Food Profile Kylie R. Jeffrey and Caroline N. Rivera
| Variable | Kit Sent (n=109 adults) | Kit Returned (n=79 adults) |
| --- | --- | --- |
| Other | 18 (16.5%) | 12 (15.2%) |
| **Meal Preparation & Food Decisions (past 30 days)** |  |  |
| Subscribed to meal prep service | 5 (4.6%) | 3 (3.8%) |
| **Who primarily decided what you ate** |  |  |
| I made most of the decisions | 95 (87.2%) | 70 (88.6%) |
| My family or household members decided | 13 (11.9%) | 9 (11.4%) |
| The cafeteria or institutional setting decided | 1 (0.9%) | -- |
| **Who primarily prepared your food** |  |  |
| I cooked most of my food | 80 (73.4%) | 62 (78.5%) |
| Someone else in my household cooked most of my food | 18 (16.5%) | 12 (15.2%) |
| I got most of my food from restaurants or takeout | 5 (4.6%) | 1 (1.3%) |
| Other | 5 (4.6%) | 4 (5.1%) |
| I got most of my food from a cafeteria | 1 (0.9%) | -- |
| **Health Condition Impacting Food** |  |  |
| No | 93 (85.3%) | 67 (84.8%) |
| Yes | 16 (14.7%) | 12 (15.2%) |
| **Food Intolerances and Allergies** |  |  |
| Lactose Intolerance | 5 (4.6%) | 4 (5.1%) |
| Gluten Intolerance | 1 (0.9%) | 1 (1.3%) |
| Any known food allergy | 17 (15.6%) | 13 (16.5%) |
| **Supplement Use** |  |  |
| Probiotic supplement (past month) | 27 (24.8%) | 19 (24.1%) |
| Fiber supplement (past month) | 18 (16.5%) | 13 (16.5%) |
| **Antibiotic Use** |  |  |
| Yes | 40 (36.7%) | 33 (41.8%) |
| No | 68 (62.4%) | 46 (58.2%) |
| Prefer not to answer | 1 (0.9%) | -- |
| **GI Health History** |  |  |
| IBS or IBD history (current or past) | 8 (7.3%) | 4 (5.1%) |
| Anti-diarrheal medication used | 8 (7.3%) | 4 (5.1%) |
| **Bowel movement frequency (per day)** |  |  |
| Less than one | 9 (8.3%) | 9 (11.4%) |
| One | 66 (60.6%) | 49 (62.0%) |
| Two | 25 (22.9%) | 16 (20.3%) |
| Three | 8 (7.3%) | 4 (5.1%) |

Returning Genomic Dietary Data to Participants by Translating Stool DNA into a Personal Food Profile Kylie R. Jeffrey and Caroline N. Rivera
| Variable | Kit Sent (n=109 adults) | Kit Returned (n=79 adults) |
| --- | --- | --- |
| Four or more | 1 (0.9%) | 1 (1.3%) |
| Stool type (Bristol Stool Scale) |  |  |
| Constipated (Type 1-2) | 11 (10.1%) | 11 (13.9%) |
| Normal formed stool (Type 3-4) | 87 (79.8%) | 62 (78.5%) |
| Loose/watery stool (Type 5-7) | 11 (10.1%) | 6 (7.6%) |
| GI Symptoms (past 30 days, multi-select) |  |  |
| Constipation | 45 (41.3%) | 30 (38.0%) |
| Nausea | 16 (14.7%) | 12 (15.2%) |
| Vomiting | 3 (2.8%) | 3 (3.8%) |
| Diarrhea | 47 (43.1%) | 27 (34.2%) |
| Indigestion | 21 (19.3%) | 15 (19.0%) |
| Food poisoning | 3 (2.8%) | 1 (1.3%) |
| Gastroenteritis | 1 (0.9%) | -- |
| Other | 3 (2.8%) | 3 (3.8%) |
| Defecation discomfort frequency (past 30 days) |  |  |
| 0 times | 55 (50.5%) | 46 (58.2%) |
| 1–3 times | 33 (30.3%) | 21 (26.6%) |
| 4–6 times | 11 (10.1%) | 7 (8.9%) |
| 7–10 times | 5 (4.6%) | 3 (3.8%) |
| More than 10 times | 4 (3.7%) | 2 (2.5%) |

Values shown as n (%) unless otherwise noted. Multi-select items (Specialized Diets, Regular Beverage Consumption, Food Purchasing Locations, GI Symptoms) may sum to >100%. "--" indicates zero respondents. GI symptoms reflect the 30 days prior to baseline survey completion.

**Table 3.**
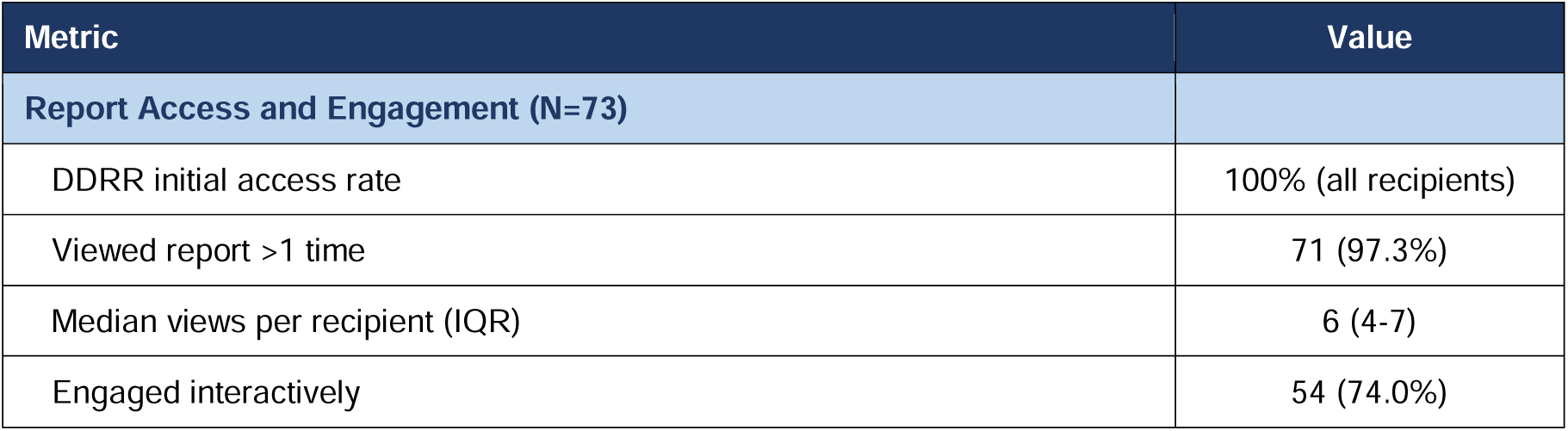
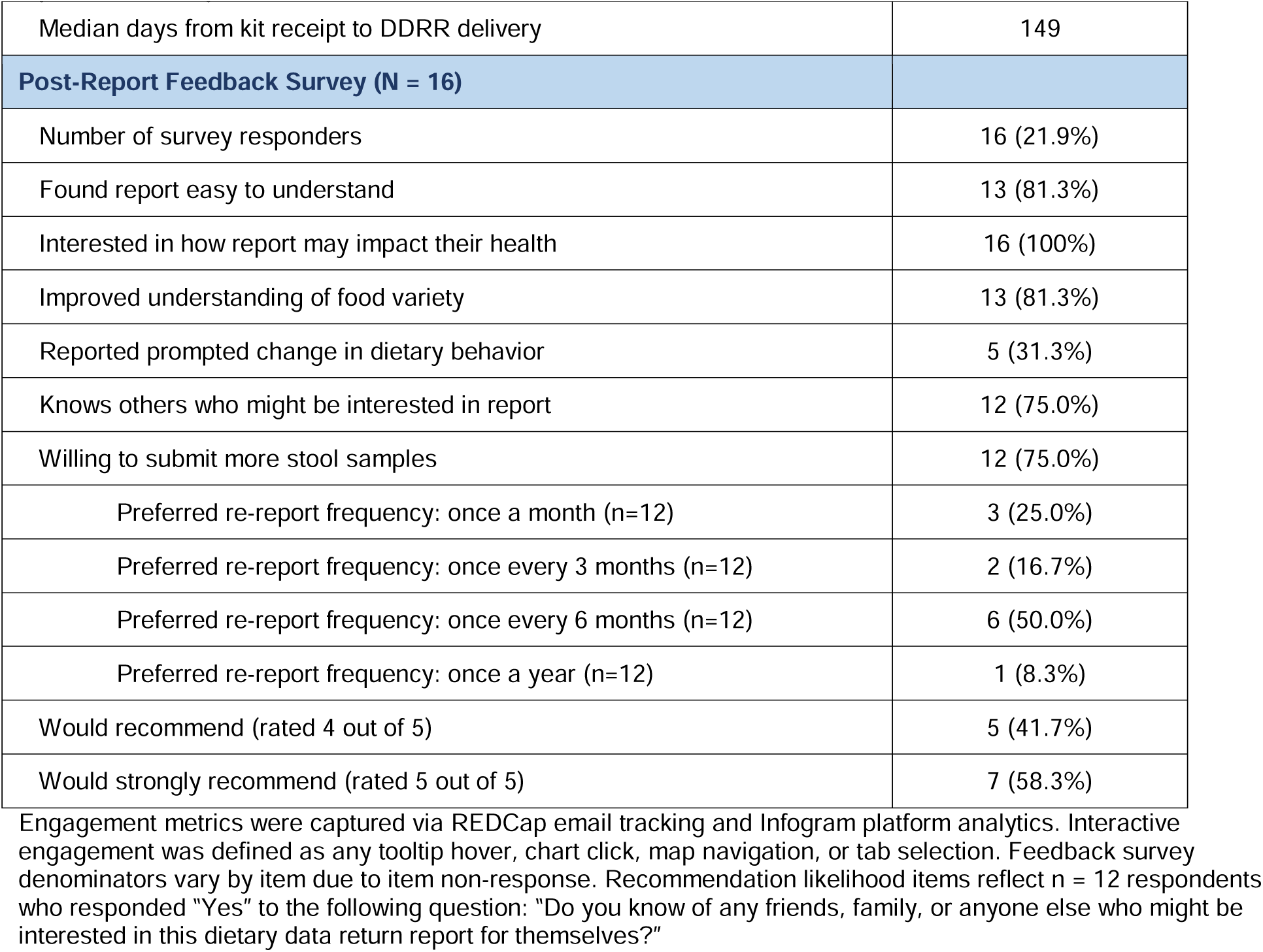
Diet Data Return Report engagement metrics and post-report feedback survey results. Engagement with the personalized DDRR among recipients with available analytics data (n=73), and responses to the post-report feedback survey among completers (n=16, 21.9% of DDRR recipients).

### 3.4 FoodSeq Produced Coherent, Recognizable Dietary Data

All 80 returned samples passed the minimum read-depth threshold for the trnL marker. One sample failed the 12SV5 read-depth threshold and was excluded from downstream animal diversity analyses (**Supplemental Figure 1**). FoodSeq detected 121 food taxa across all eight food groups cohort-wide, including 28 vegetable, 23 fruit, 13 seed and nut, 13 herb and spice, 11 legume, 8 meat and poultry, 7 grain, and 5 seafood taxa (**Supplemental Figure 2A**). Cohort-wide pMR ranged from 6 to 32, with a median of 17 distinct plant taxa per sample (**Supplemental Figure 2C**). Among the 79 samples with successful 12SV5 sequencing, aMR ranged from 0 to 6, with a median of 2 animal taxa per sample (**Supplemental Figure 2D**).

Self-reported diet was predominantly omnivorous (n = 66; 83.5%), with smaller pescatarian (10.1%), vegetarian (3.8%), and vegan (2.5%) groups, and most participants (75.5%) reported no specialized dietary pattern (**Table 2**). aMR was consistent with self-reported diet type: omnivores showed the broadest range of animal-derived detections, while vegetarian and vegan participants had no detectable animal-derived DNA (**Supplemental Figure 2B**).

### 3.5 Report Comprehension and Acceptability

Among the 16 participants who completed the feedback survey, the report was broadly understood and well received (**Figure 4**). Thirteen of 16 (81.2%) found the report easy to understand, 13/16 (81.2%) reported improved understanding of their dietary variety, and all 16 (100%) expressed interest in how their results might relate to their health. Five respondents (31.2%) reported that the report prompted a dietary behavior change, most often increasing fruit or vegetable consumption or changing food purchasing habits. Future participation intent was high, with 12/16 (75%) willing to submit additional samples and 12/16 (75%) reporting that they knew others who might want a similar report; all 12 respondents who rated recommendation likelihood gave a 4 or 5 out of 5. Open-text comments echoed these themes, with participants describing new dietary self-reflection (“I was kind of surprised that my meat and seafood intake was average” [P15]) and easy comprehension (“Very easy to comprehend (and easy on the eyes too!)” [P7]), while the most common complaint was turnaround time (“I would not do this again right now because the response time was way too long” [P6]).

When participants questioned their report, their comments concentrated at the boundary between sequence and food. Half of respondents (8/16) named at least one food they eat often that was absent from their report, and the categories they cited most were herbs and spices, seafood, and fruits. A smaller group (3/16) reported detections they had not expected, offering examples such as “green peas, chamomile, sesame” and “bananas/plantains”. Both patterns involve foods that are easy to overlook in self-report, and, in several cases, foods present in small amounts or in processed form, where DNA recovery is lower and a single marker sequence may map to more than one food. The feedback sample is small and these observations are directional, but the clustering of participant-flagged discrepancies at the interface between sequence and food points to interpretation, rather than sequencing, as the step that most shapes how these data are received.

## 4. Discussion

### 4.1 Principal Findings

Everyone EATS shows that genomic dietary data can be returned to participants. We translated FoodSeq output into a personalized report, delivered it without payment, and participants engaged with it. A 72.1% kit return rate and high sequencing success rate established the operational viability of the pipeline in a remote setting, and among participants who received a DDRR, engagement was high, with all accessing their report, 97% viewing it more than once, and 74% engaging interactively with report content. These engagement levels are notable given that no financial compensation was offered and the median interval from sample collection to report delivery was 149 days. Among the 16 participants who completed the post-report feedback survey, 81.2% found the report easy to understand, 81.2% reported improved understanding of their dietary variety, and 31.2% reported a dietary behavior change.

### 4.2 Curiosity as a Participation Driver and Its Equity Implications

The Personal Genome Project, in which thousands of participants enrolled without compensation in exchange for receiving personal genome sequences, provides the clearest precedent for data return as a participation incentive (24). Everyone EATS tested whether a similar model might work for stool-based genomic dietary data. With no financial compensation offered, 79.8% of participants cited curiosity about their personal dietary data and/or cited support for broader research goals as a primary motivation for enrolling. That 80 of 111 mailed kits were returned under these conditions, and that 75% of feedback completers expressed willingness to submit additional samples, suggests that interest in molecular self-knowledge can sustain participation across multiple contacts. These findings extend a well-documented pattern of motivation for personal biological data across microbiome (12), direct-to-consumer (25, 26), and personalized genomic contexts (27–29) into dietary metabarcoding.

Of note, the Everyone EATS cohort was predominantly white, female, and well-educated, an early-adopter profile consistent with the demographic profiles reported in American Gut and personalized genomics research (29, 30). These data suggest curiosity-driven recruitment should not be considered broadly sufficient; the populations with the most to gain from precision nutrition tools, including those facing diet-related chronic disease disparities, are unlikely to be reached through the low-intensity recruitment model used here. What these data do establish, however, is a proof of concept that genomic dietary data return can function as a genuine participant incentive in at least one population, and the design infrastructure developed here (interactive reports, food-group translation, population contextualization) provides a foundation to build from. Extending that foundation equitably, through community-partnered recruitment, multilingual design, and compensation models appropriate for underserved populations, will be an important future step.

### 4.3 The Interpretation Gap: Analytical Limits and Participant Recall

The foods participants flagged as missing or unexpected were not distributed randomly across the diet, and they point to where sequence-to-food translation is most acute. Reported omissions concentrated among herbs and spices, seafood, and fruits, while unexpected detections included items such as green peas, chamomile, and sesame. Discordance of this kind, between what a FoodSeq report shows and what a participant expects, can arise from either the analytical or participant side.

On the method side, FoodSeq has known recovery limits. Prior work suggests that select highly processed or diluted items such as granulated sugar and coffee leave little residual DNA to detect (10). Yet, on the participant side, memory is also imperfect (31–33). Self-report routinely misses foods eaten in small amounts, ingredients folded into mixed or processed dishes, and components a person never knew were present (32). Foods like sesame, garlic, and chamomile can hide in oils, sauces, blends, and teas, so an unexpected detection could be a real food the participant did not recall rather than a spurious signal (10).

Assessing FoodSeq accuracy requires gold-standard dietary records not collected in this pilot. As the field works toward orchestrating complementary dietary-assessment tools (34), building and validating this translation layer, and conveying its uncertainty to participants, are the prerequisite for returning genomic dietary data at scale.

### 4.4 Species-Level Resolution as a Novel Frame for Dietary Communication

The same property that makes sequence-to-food translation technically demanding, species-level resolution, is also what distinguishes these data from anything else in nutrition practice or consumer-facing dietary products. Where conventional dietary assessment asks how much protein, fiber, or macronutrients a person consumed, FoodSeq asks which plant and animal species were present in their stool, and in what combination. Research in food biodiversity has established dietary species richness as a meaningful axis of nutritional quality: each additional food species consumed is associated with improved micronutrient adequacy across diverse populations (35), and higher dietary species richness tracks with adherence to dietary guidelines in high-income settings (36). To date, dietary species richness has been measured exclusively through self-report. FoodSeq offers an objective measure of the same construct from biological specimens. That participants who reported improved understanding of their food variety described the experience as personally surprising points to species-level profiles engaging people through a different register than conventional dietary feedback. Whether that translates into sustained behavior change is an empirical question this pilot cannot answer. What these data do suggest is that precision nutrition may benefit from diversifying the language in which dietary data is communicated; macronutrients, calories, and clinical risk scores may be only one of several registers in which dietary information can reach people.

### 4.5 Design Principles for Genomic Dietary Data Return

The Everyone EATS pilot, viewed alongside the return-of-results literature, points toward several practical principles for designing genomic dietary data return. We offer these as provisional guidelines derived from a small pilot, not as a prescriptive standard.

- **Translation** – Participants should not be asked to interpret raw molecular outputs. The DDRR organized detected food taxa into familiar food groups and anchored the primary diversity metric in population comparisons from prior published FoodSeq studies. The 81.2% comprehension rate suggests the food-group framing translated molecular outputs into terms participants could evaluate. Future designs should test alternative translation strategies, including nutrient-adjacent summaries, food systems narratives, and temporal comparisons across repeated samples, and evaluate these strategies explicitly in populations with varying health and food literacy.
- **Population contextualization** – A pMR value is interpretable in isolation only for readers already familiar with the metric; positioned against a distribution of cohort benchmarks, it becomes a legible self-assessment. This principle is well-established in genetic testing (16) and microbiome data return (12, 37), and appears to generalize to dietary metabarcoding. Importantly, reference populations should reflect the food cultural and socioeconomic diversity of the target participant population. The microbiome field offers a cautionary precedent (37, 38): cohorts assembled from high-income Western settings encode diversity norms that may systematically position participants from other cultural or economic contexts as outliers, not because their diets lack diversity but because that diversity is organized differently (35).
- **Interactive delivery** – The 74% interactive engagement rate and a median of six report views per recipient are consistent with evidence from digital health research that interactive formats support comprehension and sustained reference beyond initial delivery (39). Static reports delivered as PDFs are unlikely to achieve comparable engagement with a data type as unfamiliar as dietary metabarcoding.
- **Turnaround time** – This is the design constraint most directly within a research team’s control, and the one most in need of improvement. The median 149-day interval from sample receipt to report delivery was the most commonly raised concern in participant open-text feedback and almost certainly contributed to the low feedback survey completion rate. For data return to function as a genuine participant engagement strategy, particularly in longitudinal designs, reports must be delivered close enough in time to the sample contribution to feel connected to it. Automated pipeline triggering, replacing the manual batch approach used in this pilot, is a prerequisite for scaled deployment, and a target of four to six weeks from sample receipt to report delivery is achievable with current infrastructure.
- **Honest communication** – Explanation of data limitations is essential for participant trust. The DDRR embedded method-specific caveats explaining slash-grouped taxa, incomplete detection, and the absence of mass quantification as expected features of DNA-based food tracking rather than analytical errors. The study website also contained a dedicated Frequently Asked Questions page (**Supplementary Material**). Report gaps and unexpected detections should be explained within the report as expected features of how the method works, not relegated to a disclaimer.

### 4.6 Limitations

Several limitations constrain the interpretation of these findings. The Everyone EATS cohort was predominantly white (85.3%), female (78.0%), and well-educated (71.6% graduate degree), with relatively low rates of food insecurity, so comprehension and engagement rates observed here may substantially overestimate what would be found in more socioeconomically diverse populations or in those with lower health or food literacy. If genomic dietary data return is deployed at scale without equity-centered design, it risks becoming a precision nutrition tool that serves those already advantaged in dietary knowledge and health system access. The sequence-to-food rules used here also encode population-specific assumptions about which foods are most likely eaten and in what quantity, and they are unlikely to transfer cleanly to other cuisines or populations without revalidation. The feedback survey sample (n = 16, 21.9% of DDRR recipients) is small and subject to selection bias, since respondents are a self-selected subset of an already self-selected cohort, likely skewed toward those most engaged with their results. The low final feedback survey completion rate reflects a design limitation as well; survey fatigue across a length study timeline, compounded by the 149-day delay, likely attenuated respondent motivation and contributed to non-random attrition. The dietary behavior changes reported by 31.2% of feedback completers cannot be attributed to the data return itself. Everyone EATS included no control condition, and observed changes may reflect general health motivation among a self-selected population, novelty effects, or behavioral awareness from participation rather than a causal effect of the personalized report. Finally, the sustainability of curiosity-driven enrollment without financial compensation is untested beyond a single pilot contact. Whether comparable motivation persists across repeated sampling cycles, or extends to populations less familiar with genomic research, remains an open empirical question.

## 5. Conclusions

The Everyone EATS pilot shows that genomic dietary data can be returned to participants in a form they find comprehensible and engaging, and that curiosity about one’s own diet can motivate participation. Nevertheless, realizing this approach will require a validated sequence-to-food translation layer, more diverse cohorts, evidence on the contribution of personalization, and faster turnaround between sample and report. Sequencing is not the bottleneck but rather building and validating the translational infrastructure that is meaningful across the full diversity of human diets.

## Supporting information

Supplementary Material - Sample DDRR

Supplementary Material - DDRR FAQs

Supplementary Material - Feedback Survey

Supplementary Material - Baseline Survey

Supplementary Material - STROBE-nut checklist

## Author Contributions

KRJ, CNR, MCK, AEB, and LAD designed research; KRJ, CNR, AA, and MJG conceptualized the data return; KRJ, CNR, NWI, and SJ conducted research; KRJ and CNR analyzed data; KRJ, CNR, and LAD wrote the paper. LAD had primary responsibility for final content. All authors read and approved the final manuscript.

## Acknowledgements

We thank the participants of Everyone EATS for their time and contributions. We thank Nicole Duncan for visual design and content development across the Everyone EATS study; Sana Aqeel for design assistance on the Diet Data Return Report; and Olivia Vaz for logistical study support. We thank the Forever Learning Institute in the Duke University Alumni Engagement and Development Office for their support with participant recruitment.

## Data Availability

Data described in the manuscript, code book, and analytic code will be made available upon reasonable request pending PI approval, given that data include personally sensitive participant information. The bioinformatic pipeline used to process FoodSeq data is publicly available at https://doi.org/10.5281/zenodo.20174235.

## Declaration of Generative AI

AI and AI-assisted technologies in the writing process: During the preparation of this work the author(s) used Claude Sonnet 4.6 and Opus 4.8 (Anthropic) in order to assist data analysis and draft and revise manuscript text. After using this tool, the authors reviewed and edited the content as needed and take full responsibility for the content of the publication.

