## Supplementary Material - Sample DDRR for "Returning Genomic Dietary Data to Participants by Translating Stool DNA into a Personal Food Profile"

### everyone EATS

#### Participant ID: 12345

#### How it works

1 **COLLECTION**
You send us poop samples

2 **SEQUENCING**
We sequence the food DNA from your sample

3 **RESULTS**
You receive your test results

#### What we do

In our lab, we study **how the food you eat affects your health**—from your gut microbiome to your energy levels, digestion, and beyond. We collect stool samples to find traces of dietary DNA—yes, food has DNA, too! This approach lets us see not just what you remember eating, but **what your body actually consumed**.

#### Why it matters

Your **diet plays a powerful role** in how you feel and function, but historically, it's been challenging to accurately measure real-world eating habits. Our research leverages **DNA-based food tracking** to help fill that gap. By understanding these connections, we can uncover patterns that support **evidence-based dietary choices**—for individuals and for communities.

**REMEMBER**: This report is for personal information use only, not for diagnostic or forensic purposes. The DNA analysis methods used do not measure food mass, which is important for health assessment. We are not a certified clinical diagnostic lab.

### Your Food Patterns

#### What did we find in your poop?

Based on the DNA in your poop, we detected a variety of items from different food groups:

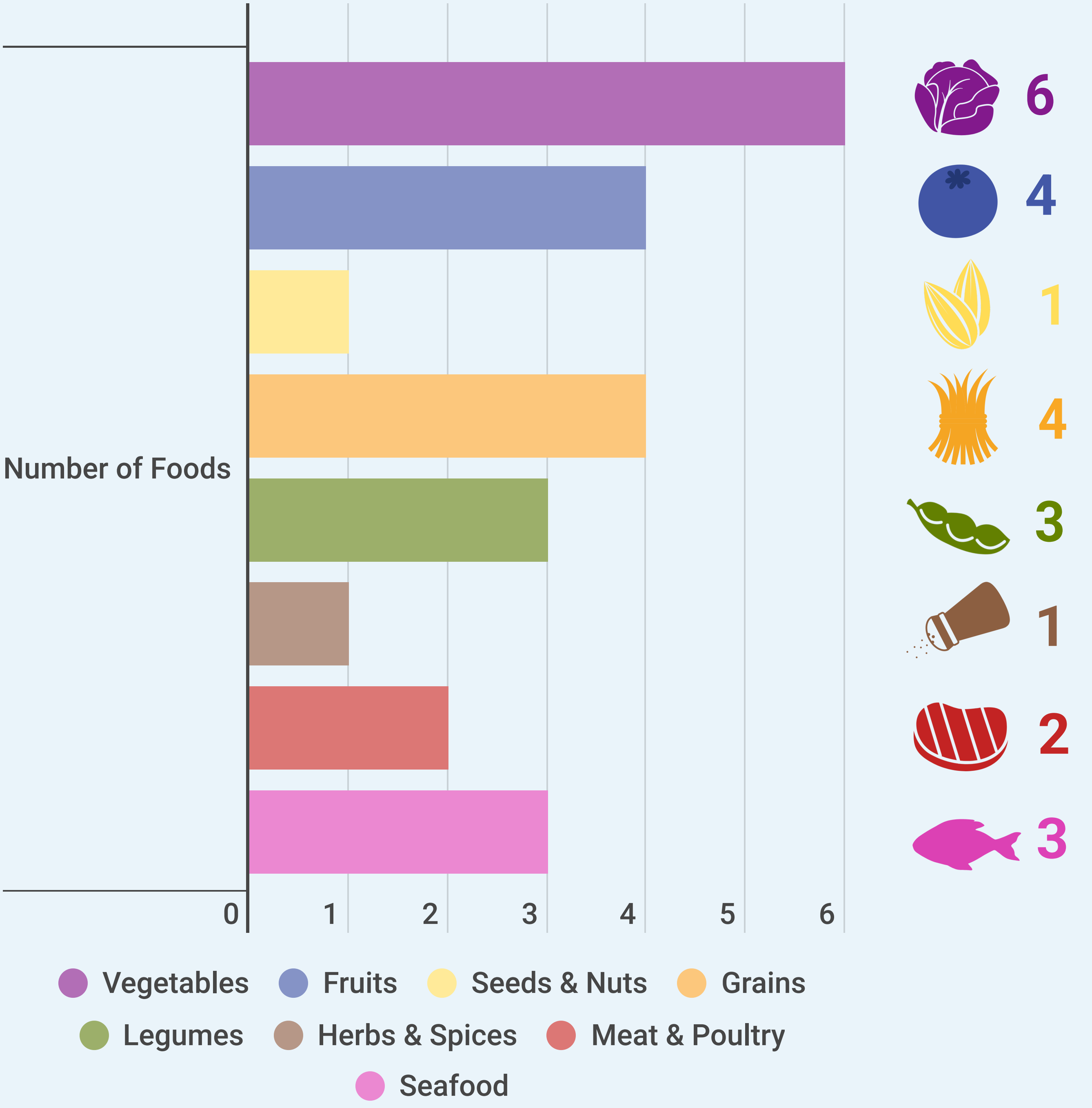


**REMEMBER**: This report is for personal information use only, not for diagnostic or forensic purposes. The DNA analysis methods used do not measure food mass, which is important for health assessment. We are not a certified clinical diagnostic lab.

### Your Food Patterns

#### What specific foods did we find?

Right now, we can only detect DNA from plants and certain animals. In some cases, we can't tell apart foods by their DNA, so we write these foods separated with a slash, like "apples/pears". Similarly, because they both come from cows, we can't distinguish beef and dairy products. Sometimes we detect different DNA for the same food so we list that food again.

| Food Group | Food Detected |
|---|---|
| Vegetables | Okra |
| | Lettuce |
| | Spinach |
| | Cabbage family |
| | Asparagus |
| | Carrot / Parsley / Cilantro / Dill / Fennel / Cumin |
| Fruits | Banana |
| | Grapes |
| | Blueberry |
| | Strawberries / Rose / Dewberry / Cloudberry / Raspberry / Wineberry / Blackberry |
| Seeds & Nuts | Flaxseed |
| Grains | Oat |
| | Wheat |
| | Corn |
| | Rice |
| Legumes | Soy |
| | Fenugreek |
| Herbs & Spices | Black pepper |
| Meat & Poultry | Beef / Dairy |
| | Chicken / Eggs |
| Seafood | Atlantic salmon |
| | Mahi-mahi |
| | Black sea bass |
| Other | Chocolate |
| | Tea |

### Dietary Diversity in the U.S.

#### What is Dietary Diversity?

Dietary diversity refers to the variety of different foods and food groups consumed. Research shows that people who eat a wider variety of foods tend to have better nutrition and health outcomes.

#### Did you know?

One way the Dietary Guidelines for Americans assess dietary quality is by measuring variety within and across food groups. Recent reports have found:

**80%** More than 80% of Americans don't meet recommended intake for fruits or vegetables

**50%** More than 50% of Americans exceed recommendations for total grain and protein intake BUT don't meet subgroup totals (like whole grains and seafood)

**10%** Only 10% of Americans meet recommendations for vegetable intake

**Using our sequencing technology, we've analyzed samples from people around the United States with diverse dietary patterns, counting the number of different food items detected from each food group. Here's how your sample compares to the average:**

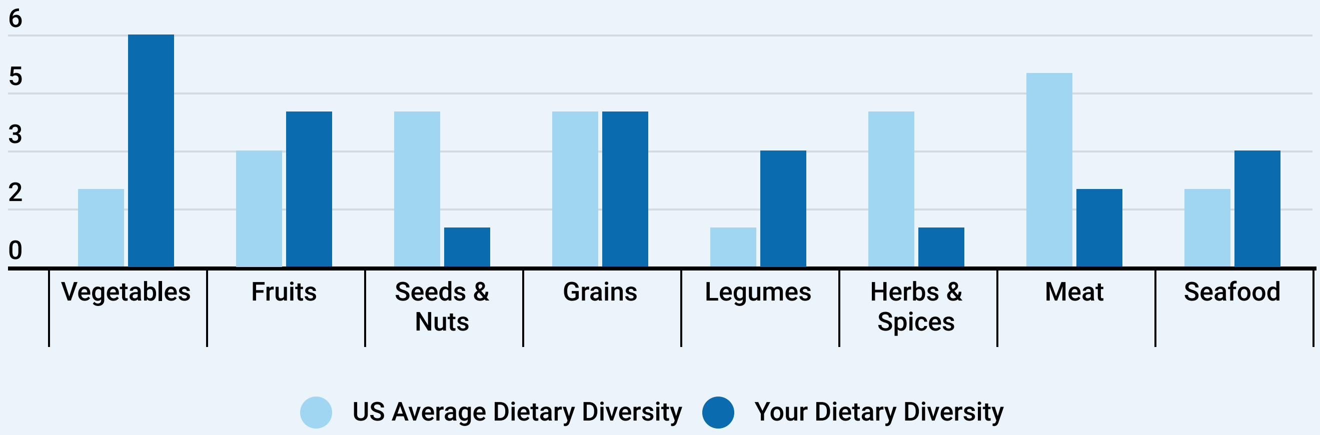


**REMEMBER**: This report is for personal information use only, not for diagnostic or forensic purposes. The DNA analysis methods used do not measure food mass, which is important for health assessment. We are not a certified clinical diagnostic lab.

### Dietary Diversity in the U.S.

#### Why Variety Matters

The Dietary Guidelines for Americans recommend 2-3 cups of vegetables per day, distributed across five subgroups. Each subgroup provides unique nutrients your body needs:

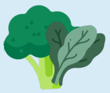

**Dark Greens vegetables** like spinach, broccoli, and kale are packed with folate, vitamin K, iron, and calcium that support blood health, bone strength, and eye health.

**Red & Orange vegetables** such as carrots and red peppers are rich in vitamin A, vitamin C, and potassium—essential for healthy vision, immune function, and normal blood pressure.

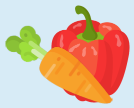

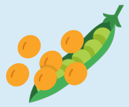

**Beans, peas, & lentils** including black beans, chickpeas, and lentils provide plant-based protein, fiber, folate, and iron that support digestive health and steady energy.

**Starchy vegetables** like potatoes, corn, and winter squash provide complex carbohydrates, B vitamins, potassium, and fiber for sustained energy and vital functions.

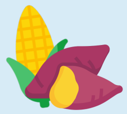

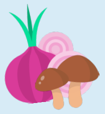

**Other vegetables** including mushrooms, onions, and cauliflower bring diverse benefits from immune-supporting compounds to vitamin D and protective plant nutrients.

**We measured the average number of different fruit and vegetable subgroups** (described above) **detected across American samples. Here's how yours compares:**

| Your Dietary Diversity | US Average Dietary Diversity |
|---|---|

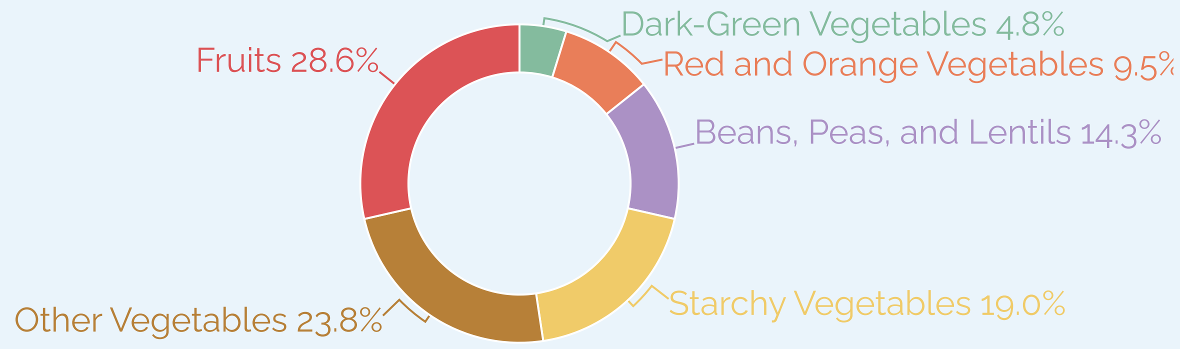


**REMEMBER**: This report is for personal information use only, not for diagnostic or forensic purposes. The DNA analysis methods used do not measure food mass, which is important for health assessment. We are not a certified clinical diagnostic lab.

### Dietary Diversity in the U.S.

#### What Can You Do?

Meeting dietary guidelines can feel overwhelming—tracking subgroups and cup measurements isn't always practical in daily life.

**One approach:** "Eating the rainbow." Research suggests that focusing on colorful foods may be a simpler way to achieve dietary variety. Different colors signal different nutrients, so a colorful plate often naturally includes multiple vegetable subgroups. Many nutrition educators use this color-based framework as an accessible alternative to tracking specific categories.

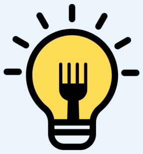

**Did you know...**

The compounds that give fruits and vegetables their colors can provide health benefits? For example, foods like tomatoes get their red color from lycopene, a powerful antioxidant that can help lower bad cholesterol.

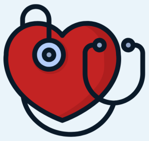

**These benefits include:**

- Decreased risk of developing type 2 diabetes
- Decreased risk of cardiovascular disease
- Some protection against certain cancers

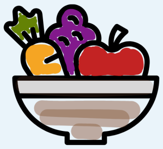

**BUT no single colored compound can do it all**

It's important to eat a variety of fruits and veggies for optimal health and nutrition. Not sure you're getting the right balance? Just follow the rainbow!

**Here's the fruit & vegetable rainbow that we've observed across all our American participants and how it compares to your food rainbow:**

Your Food Rainbow | Average USA Food Rainbow

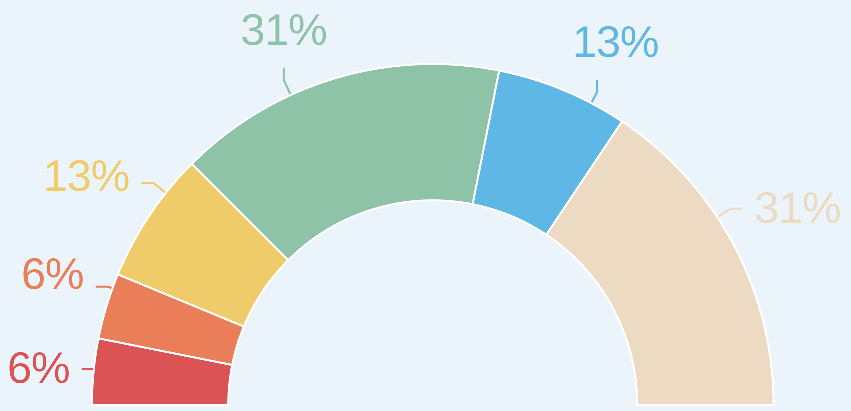


**REMEMBER**: This report is for personal information use only, not for diagnostic or forensic purposes. The DNA analysis methods used do not measure food mass, which is important for health assessment. We are not a certified clinical diagnostic lab.

### Food Processing

#### What is Food Processing?

Food processing ranges from simple steps like washing and cutting to complex manufacturing using refined ingredients such as starches, oils, and additives. Studying this spectrum helps scientists understand how different foods appear in our diets and how they may relate to health.

#### What We Found

Using FoodSeq, we identified a distinct "processing signature" in dietary DNA driven primarily by the frequent presence of three ingredients: **soy, corn, and wheat**. Their industrial derivatives (corn syrup, soybean oil, modified starches) appear widely in processed foods. This signature correlates with health measures like BMI and cardiovascular risk markers. Importantly, this doesn't mean these crops are unhealthy—in whole forms, they're nutritious. The signature reflects how their processed forms appear across many packaged foods.

#### Your Score and What it Means

Your score shows how closely your dietary pattern aligns with this processing signature on a scale from the least to most processed patterns we've observed in our sample population:

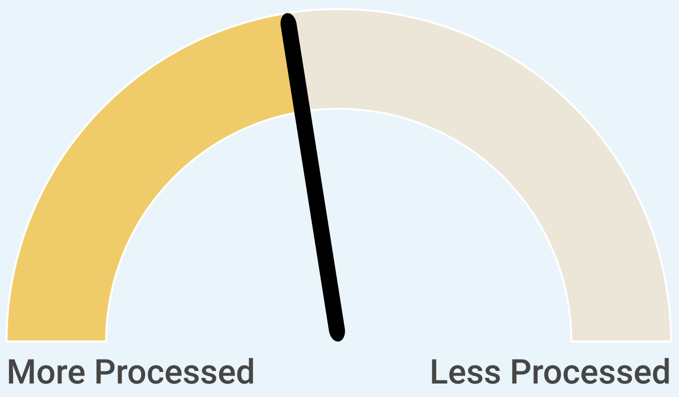


This score provides awareness, not judgment. Many factors influence what we eat, including cost, access, culture, and convenience. Research on food processing is evolving, and this signature represents one approach to understanding dietary patterns. Small, realistic shifts like cooking one more meal at home can influence patterns over time.

**REMEMBER**: This report is for personal information use only, not for diagnostic or forensic purposes. The DNA analysis methods used do not measure food mass, which is important for health assessment. We are not a certified clinical diagnostic lab.

### Thank You!

Your participation in this project has been so valuable in helping us develop the most helpful tool for people like you. We value your feedback and would love to hear from you!

#### Here are a few ways you can help:

Take a **short online survey** about your experiences with the data return. It takes less than 10 minutes!

Contact us to schedule a **brief phone/video interview**. The call will only take 30-45 minutes, and we can work around your schedule! You can contact us at **** or **919-684-8712**.
