## Supplementary Material - DDRR FAQs for "Returning Genomic Dietary Data to Participants by Translating Stool DNA into a Personal Food Profile"

*The following FAQs may be provided to interested individuals before consent through the lab website, recruitment, etc. and in the diet data return report packet.*

**General**

- **Why should I be interested in the information in my report?**
  *It contains objective information on the foods you've eaten. This contrasts with the current methods of collecting dietary information from participants which rely on subjective information from a survey or questionnaire. We're also able to capture foods often missed by these surveys, like traditional and culturally relevant ingredients.*
- **Does the report show all the foods that I ate in one day?**
  *This really depends on the person, as digestion is very individualized. In some cases, the foods we find can be reflective of just one day if someone has a fast digestive system. But it's entirely possible that what we find could range from 1 – 5 days before sample collection.*
- **What if I see foods that I know I DIDN'T eat?**
  *There are a few reasons for this:*
  *Our method is still under development, so while it's mostly very accurate, it's not perfect. Sometimes DNA from dust or another sample may get into your sample and be detected. Alternatively, foods can sometimes have surprising ingredients. For example, corn, soy, and palm oil are added to processed foods in form of syrups, extracts, or oils.*
- **Why can't I see foods that I know I DID eat?**
  *Sometimes the DNA in food will not survive long enough to be detected in stool. Generally, we find that the more someone has eaten a food, and the less processed it is, the more likely we are to see it. Additionally, we're still ironing out a few kinks to improve accuracy in our detection and reporting of fungi (for example, mushrooms) and invertebrates (for example, shrimp, clam, snails, edible bugs), so you may not see these in your report.*
- **Can you tell me the quantities of food I ate?**
  *No, but we're working on this.*
- **Should I eat differently while participating?**
  *There's no need to eat any differently. In fact, the closer you keep to your regular diet, the more reflective your report will be of your usual food preferences. This is especially useful to researchers like us who are trying to establish any relationships between what people are eating and how this affects health.*
- **Can you tell me if I have any food allergies?**
  *No, we are not able to provide any information on food allergies with this technique.*
- **Can you provide health advice based on my report?**
  *No, we do not provide health advice as we are not qualified to do so. This is the role of a registered dietician and other medical professionals. Our hope is that these Dietary Data Return Reports will be useful to these kinds of medical professionals in providing accurate, objective information on people's dietary diversity, so they know how best to help them improve their help.*

**Eligibility**

- **Can I participate if I have a health condition?**

*Yes, anyone is welcome to participate. If you are taking antibiotics, we ask that you wait to provide a sample until one month after your antibiotic course is complete.*

- **Does it cost money to participate and receive a report?**
  *There is no cost to you.*
- **Where is the study taking place?**
  *Our lab, based at Duke University in Durham, North Carolina, has primarily focused its studies within the Durham area, meaning most individuals who have received the Dietary Data Return Report currently reside nearby. However, we have collected thousands of samples from across the globe, enabling us to compare your data to global food patterns. While logistics, such as shipping stool samples, have influenced the concentration of return reports in the Durham area, our goal is to expand access to the reports across the United States and eventually worldwide.*

**Sampling**

- **How do I provide a stool sample?**
  *We will provide a sample collection kit with clear instructions on how to sample your stool, store it safely, and then ship it.*
- **What happens if I lose my sample kit?**
- **How quickly must I ship the sample?**
  *Within 48 hours. It can be stored in your personal freezer until you are able to ship it.*

**Privacy**

- **Do I need to share personal information about myself?**
  *We might ask for basic demographic (i.e age, race, gender), anthropometric (i.e height, weight), and lifestyle behavior (i.e level of physical activity) information, depending on the study. You are under no obligation to answer these questions if you don't want to.*
- **Can I withdraw from the study?**
  *Yes, you are free to withdraw from the study at any time.*
- **If I withdraw, what will happen to my information and my stool sample?**
  *Should you withdraw, any and all study information, along with your stool sample, will be destroyed.*
- **What are the risks of participating in this study?**
  *The only risks in this study are possible risk of infection while sampling stool and possible loss of confidentiality. Every effort will be made to ensure all information is kept strictly confidential.*
- **Is my genetic information being used for this study?**
  *No. We are not performing any experiments involving your DNA. We are looking for the DNA found in the plant foods that you consumed.*
- **How will my privacy be protected?**
  *Your privacy is a top priority. Every effort will be made to keep your information confidential, though absolute privacy cannot be guaranteed. Identifying details will only be shared when necessary for the study and will be removed when possible. While your data may be*

*reviewed by regulatory groups or included in scientific presentations, your identity will not be disclosed, and samples will be labeled with a unique ID to protect your confidentiality.*

**Feedback**

- **Where and how do I provide feedback?**
  *A link is provided on your Dietary Data Return Report that will take you to a survey in which you can provide feedback. In addition, if you'd like to schedule a phone or video interview, you can reach out to the study team directly at.*
