## Supplementary Material - Feedback Survey for "Returning Genomic Dietary Data to Participants by Translating Stool DNA into a Personal Food Profile"

Diet Data Return Report Feedback Survey

1. Was this report easy to understand?
    a. Yes
    b. No

2. Do you have any suggestions for how we can make this report easier to understand?
   *[Free form response]*

3. Did reading the report improve your understanding of the variety of foods in your diet?
    a. Yes
    b. No

4. Was there anything you eat often, that was missing from the report?
   *[Free form response]*

5. Was there anything in this report that you thought you didn't eat?
   *[Free form response]*

6. Is there anything else you found surprising about the foods in your report?
   *[Free form response]*

7. Has the information in your report prompted you to pursue any changes in your behavior?
    a. Yes
    b. No

8. If yes, what did it prompt you to do? Select all that apply.
    a. Talk to your friends and family about your food consumption
    b. Talk to a dietitian or other health professional
    c. Increase your fruit and/or vegetable consumption
    d. Increase your meat and/or dairy consumption
    e. Change your food purchasing habits
    f. Other

9. Do you know of any friends, family, or anyone else who might be interested in this dietary data return report for themselves?
    a. Yes
    b. No

10. If yes, how strongly would you recommend it to them?
    a. 1 – wouldn't recommend
    b. 2
    c. 3
    d. 4
    e. 5 – would definitely recommend

11. Are you interested in how the information in your diet data return report may impact your health?
    a. Yes
    b. No

12. If yes, please tell us why:
    *[Free form response]*

13. Would you consider sending in more stool samples, if it meant you could address some of the things you've described above?
    a. Yes
    b. No

14. If yes, how frequently would you like to receive a new report?
    a. Once a week
    b. Once every 2 weeks
    c. Once a month
    d. Once every 3 months
    e. Once every 6 months
    f. Once a year

15. If you had the return of results in real-time (e.g., a week after you provided the sample), would it affect your food choices? Why or why not?
    [Free form response]

16. Would you consider the information provided in the Diet Data Return Report valuable enough to pay for it as a service in the future?
    a. Yes
    b. No
    c. Maybe

17. ***If no,*** please explain what information, if any, was missing from the report that would make it valuable enough for you to consider payment (to help cover the cost of DNA sequencing and analysis).

18. ***If yes or maybe,*** what amount would you be comfortable paying for a *single* report?

19. ***If yes or maybe,*** what amount would you be comfortable paying for follow-up *repeat* reports?

20. Is there anything else you think we should know about your experience with the study?
    [Free form response]
