## Supplementary Material - Baseline Survey for "Returning Genomic Dietary Data to Participants by Translating Stool DNA into a Personal Food Profile"

Date:

**GENERAL INFORMATION**

1. Date of birth:
2. What is your current age?
3. Current Address
    a. Street address:
    b. City:
    c. State:
    d. Zip code:
4. What sex were you assigned at birth? *Female/Male/Prefer not to answer*
5. How do you currently describe your gender? *Female/Male/Transgender/Non-binary/Other/Prefer not to answer*
6. Country of birth:
    a. If outside of the US, what year did you begin residing in the US full-time:
7. Please select your race - select all that apply: *American Indian/Alaska Native/Asian/Native Hawaiian or Other Pacific Islander/ Black or African American/White/Other/Prefer not to say*
8. Are you Hispanic or Latino? *Yes/No/Prefer not to say*
9. Please describe your ethnicity in more detail (optional):
   *For example: Asian American, Laotian, Korean/Japanese, parents immigrated from Ghana, Afro-Caribbean, Ashkenazi Jewish, etc.*
10. What is your highest level of education? *Did not complete high school/High school or GED equivalent/Some college or technical school/Associate's degree/Bachelor's degree/Some graduate school or professional/ Graduate or professional degree/Prefer not to answer*
11. What is your household income? *$0 / $1 to $9 999 / $10 000 to $24 999 / $25 000 to 49 999 / $50 000 to 74 999 / $75 000 to 99 999 / $100 000 to 149 999 / $150 000 and greater / Prefer not to answer*
12. Which of this best describe your living situation? Please select all that apply: *Live in a large city, a suburb near a large city, a small city or town, or a rural area*
13. Generally speaking, are you confident that the tap water in your house is safe to drink?
    *Y/N/Prefer not to answer*

14. How far do you reside from a park? *1 block / 2 blocks / ¼ mile / ½ mile or more/Prefer not to answer*

15. During the last 12 months, was there a time when you needed healthcare but didn't get it because you couldn't afford it? *Y/N/Prefer not to answer*

16. During the last 12 months, was there a time when you didn't have enough to eat because you couldn't afford it? *Y/N/Prefer not to answer*

17. Have you used antibiotics in the last year? *Y/N/Prefer not to answer*

    a. If yes, how many times?

18. What best describes your main reason for wanting to receive a Diet Data Return Report? *Select all that apply: I am curious about my personal data/I am generally health-conscious/I have a specific health condition related to diet/I want to optimize my health and performance/I want to understand more about what my child is eating/I am curious about food consumption in elderly communities/I am a lifelong learner and want to keep up with latest advances in science and technology/I want to see how my diet compares to others (e.g., at work, home, or in my group)/I want to support the broader goals of this research so that dietary education, intervention, and access can be optimized/Other (please specify)*

**DIET INFORMATION**

19. Which of these options best describes your diet? Select one: *I eat anything with no exclusions (omnivore)/I eat anything except red meat/Vegetarian but eat fish/Vegetarian/Vegan*

20. Do you follow any of these specialized diets? Select all that apply: *Paleo or primal diet/Modified paleo diet/Raw food diet/FODMAP/other low-grain, low processed food diet/Kosher/Mediterranean/Halaal/ Exclude dairy/Exclude gluten/Exclude refined sugars/Other/I do not follow a specialized diet*

*21.* Do you have any food allergies? *Y/N*

    *a.* If yes, please describe.

*22.* In the past 30 days, where did you purchase foods? *Select all that apply: Local farmer's market/Walmart/Co-op/Compare/Publix/Dollar stores (e.g., Dollar General, Family Dollar)/Wegmans/Gas stations/Food Lion, Harris Teeter/Kroger/H-Mart/Drug stores (e.g., CVS, Walgreens)/Aldi/Trader Joe's/Fresh Market/Target/ Whole Foods/Lidl/Wholesale clubs (e.g., Costco, Sam's Club, BJ's)/Local international grocery store/Online grocery services (e.g., Amazon Fresh, Walmart Online)/ Other*

*23.* In the past 30 days, have you subscribed to a meal prep service? *Y/N*

*24.* In the past 30 days, who primarily decided what food you consumed? *I made most of the decisions/My family or household members decided/The cafeteria or institutional setting decided/Other*

25. In the past 30 days, who was primarily responsible for preparing most of the food you consumed? *I cooked most of my food/Someone else in my household cooked most of my food/I got most of my food from a cafeteria/I got most of my food from restaurants or takeout/Other*

26. Do you have a health condition that impacts your food consumption? *Y/N*

    a. If yes*, please describe.*

27. Which of the following do you consume regularly? *Select all that apply: Coffee/Tea/Alcohol/Soda/Juice/None*

28. Is there anything else you would like the study team to know about your typical dietary habits? *Y/N*

    a. If yes, please describe.

29. In the last month, have you taken a probiotic supplement (a supplement containing active/live bacterial cultures)? *Y/N*

30. In the last month, have you taken a fiber supplement? *Y/N*

**GASTROINTESTINAL INFORMATION**

31. Do you have a history or current diagnosis of irritable bowel syndrome or inflammatory bowel disease? *Y/N*

32. Do you have a history or current diagnosis of untreated colorectal cancer? *Y/N*

33. Do you have a history or current diagnosis of intestinal obstruction? *Y/N*

34. Have you had a colonoscopy within the past 30 days? *Y/N*

35. How many times do you have a bowel movement in an average day? Select one. : *Less than one / One / Two / Three / Four or more*

36. Describe the quality of your bowel movements, using the chart below for reference. *Select one: Type 1/Type 2/ Type 3/ Type 4/ Type 5/Type 6/ Type 7*

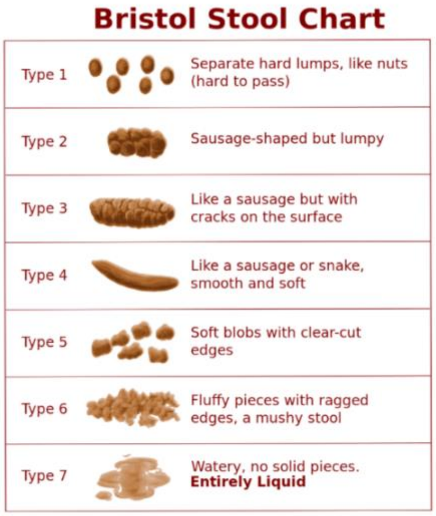


37. In the past 30 days, how many times have you experienced constipation? *0 / 1-3 / 4-6 / 7-10 / more than 10*

38. In the past 30 days, how many times have you experienced diarrhea? *0 / 1-3 / 4-6 / 7-10 / more than 10*

39. In the past 30 days, how many times have you experienced discomfort while defecating? *0 / 1-3 / 4-6 / 7-10 / more than 10*

40. In the past 30 days, have you used anti-diarrheal medications? *Y/N*

41. In the past 30 days, have you experienced any GI symptoms or illnesses? *Select all that apply: nausea/vomiting, diarrhea, indigestion, food poisoning, gastroenteritis, other*
