## Supplementary Material for "Returning Genomic Dietary Data to Participants by Translating Stool DNA into a Personal Food Profile"

**Supplementary Methods: FoodSeq Laboratory and Bioinformatic Pipeline**

**DNA Extraction**

Prior to extraction, stool samples preserved in 95% ethanol centrifuged at 3,000 rpm for 15 minutes at room temperature. The ethanol supernatant was removed, and samples were allowed to air-dry for up to 15 minutes under a biosafety cabinet to remove residual preservative. Samples were then transferred to 2-mL cryovials and stored at –20°C until extraction. Process blanks were introduced at the ethanol removal step and carried through all subsequent processing stages; one extraction blank was included for every five samples processed.

DNA was extracted from stool aliquots using the DNeasy PowerSoil Pro Kit (QIAGEN, Hilden, Germany) following the manufacturer's protocol, with the modification that the CD2 lysis incubation was performed overnight at 4°C. Samples were processed in randomized order within each batch. A 1:10 dilution was prepared in Solution C6 for downstream quantification.

DNA concentration was measured fluorimetrically using the Quant-iT dsDNA Broad Range Assay Kit (Thermo Fisher Scientific). Samples with concentrations below 10 ng/µL were re-quantified using the Quant-iT High Sensitivity kit. DNA plates were stored at –20°C prior to amplification.

**PCR Amplification and Library Preparation**

We performed FoodSeq using a two-step PCR protocol. Primary PCR amplification of multiplexed trnL and 12SV5 used the KAPA HiFI HotStart PCR kit (KAPA Biosystems) in a 10-µL volume containing 0.3µL each of 10µM forward and reverse primers (IDT) for both trnL and 12SV5, 5µL of 2X KAPA HiFi HotStart Ready Mix, 0.6µL of human blocking primer (IDT), 0.1mL of 100X SYBR Green I (Life Technologies), 0.6µL of IDTE, and 2.5µL of template DNA. The primers were trnL(UAA)g and h (1, 2) and 12SV5F and 12SV5R (3, 4)with Illumina overhang adapter sequences added at the 5’ end, and the human blocking primer was DeBarba14 HomoB (5) (**Supplemental Table 0**). Cycling conditions were an initial denaturation at 95°C for 3 min, followed by 33 cycles of 98°C for 20 s, 60°C for 15 s, and 72°C for 15 s. Each PCR batch included at least 3 positive and negative controls, and samples were only advanced to the secondary PCR if controls performed as expected (otherwise the entire batch was repeated). Secondary PCR amplification to add Illumina adapters and dual 8-bp indices for sample multiplexing was performed in a 50-µL volume containing 5µL each of 2.5 µM forward and reverse indexing primers, 25µL of 2X KAPA HiFi HotStart ReadyMix, 0.5µL of 100X SYBR Green I (Life Technologies), 9.5µL of nuclease-free water, and 5µL of primary PCR product diluted 1:100 in nuclease-free water.

Amplicons were cleaned (Ampure XP, Beckman Coulter, Brea, CA), quantified (QuantIT dsDNA assay kit, Invitrogen, Waltham, MA), and combined in equimolar ratios to create a sequencing pool. If samples could not contribute enough DNA to fully balance the pool due to low post-PCR DNA concentration, they were added up to a set volume (20 µL). Libraries were spiked with 30-50% PhiX (Illumina, San Diego, CA, USA) to mitigate low nucleotide diversity. Paired-end sequencing was carried out on an Illumina MiniSeq system according to the manufacturer’s instructions using a 300-cycle Mid kit (Illumina).

**Bioinformatic Processing**

All procedures followed the previously published FoodSeq protocol (1). Briefly, raw reads were demultiplexed using bcl2fastq v2.20.0.422, Illumina adapters were trimmed with BBDuk v38.38, and primer sequences were removed and reads trimmed to contain only the target region using cutadapt v3.4. Reads with >2 errors or quality score <2 at the first base were discarded. Remaining reads were denoised and merged to generate amplicon sequence variants (ASVs) using DADA2 v1.10.0. Taxonomic assignment was performed using the assignSpecies function in DADA2 against a curated dietary reference database (1); ASVs matching multiple taxa were assigned to the last common ancestor. For 12SV5, taxonomic assignment was done using DADA2’s assignTaxonomy function, as the 12S region in animals is far more variable than the trnL region in plants. This probabilistic, bootstrap-based approach better accommodates sequence diversity and incomplete reference coverage, avoiding the excessive unassigned reads that would result from strict exact matching (1, 3). To represent food identity rather than trnL and 12SV5 region variation, we combined ASVs assigned to the same food taxon into a single group. Plant and animal metabarcoding richness (pMR/aMR) were calculated as the count of distinct plant taxa detected per sample following previous work (1, 3). The David Lab’s full FoodSeq bioinformatic pipeline can be accessed here: [10.5281/zenodo.20174234](https://doi.org/10.5281/zenodo.20174234).

**Supplementary Table 0.** Primers used in this study. ‡ denotes 8-bp barcode sequences, available from Illumina (“Illumina Adapter Sequences,” https://support-docs.illumina.com/SHARE/AdapterSeq/illumina-adaptersequences.pdf).

| **Primer Name** | **Sequence** |
| --- | --- |
| trnL(UAA)g | TCGTCGGCAGCGTCAGATGTGTATAAGAGACAGGGGCAATCCTGAGCCA*A |
| trnL(UAA)h | GTCTCGTGGGCTCGGAGATGTGTATAAGAGACAGCCATTGAGTCTCTGCACCTAT*C |
| 12SV5F | TCGTCGGCAGCGTCAGATGTGTATAAGAGACAGTAGAACAGGCTCCTCTA*G |
| 12SV5R | GTCTCGTGGGCTCGGAGATGTGTATAAGAGACAGTTAGATACCCCACTATG*C |
| i7 indexing primer^‡^ | CAAGCAGAAGACGGCATACGAGATXXXXXXXXGTCTCGTGGGCTCGG |
| i5 indexing primer^‡^ | AATGATACGGCGACCACCGAGATCTACACXXXXXXXXTCGTCGGCAGCGTC |

**Diet Data Return Report:**

An example of the interactive DDRR can be found here: <https://infogram.com/ddrr-sample-for-website-1h984wvwx8o7d2p>

**Supplementary Table 1.** Participant characteristics by kit return status among adult participants (kit returned, n=79; non-returners, n=30). Bold rows indicate statistically significant differences (p<0.05).

| **Variable** | **Kit Returned** **(n=79)** | **Non-Returners** **(n=30)** | **p-value** | **Test** | |
| --- | --- | --- | --- | --- | --- |
| **Mean age ± SD** | **45.8 ± 16.1** | **39.8 ± 19.5** | t=0.136 **MWU=0.024** | **t-test / MWU** | |
| **Sex Assigned at Birth** | | | | | |
| **Female** | **69 (87%)** | **19 (63%)** | **0.005** | **Chi-square** | |
| **Race (multi-select)** | | | | | |
| **Asian** | **7 (9%)** | **8 (27%)** | **0.027** | **Fisher's exact** | |
| White | 70 (89%) | 23 (77%) | 0.135 | Fisher's exact | |
| **Hispanic or Latino** | | | | | |
| Yes | 2 (3%) | 2 (7%) | 0.303 | Fisher's exact | |
| **Country of Birth** | | | | | |
| United States | 69 (87%) | 25 (83%) | 0.551 | Fisher's exact | |
| Other | 10 (13%) | 5 (17%) | 0.551 | Fisher's exact | |
| **Education** | | | | | |
| Graduate/professional degree | 59 (75%) | 19 (63%) | 0.241 | Chi-square | |
| Bachelor's degree | 14 (18%) | 7 (23%) | 0.507 | Chi-square | |
| Some grad/professional school | 5 (6.3%) | 3 (10%) | 0.682 | Fisher's exact | |
| **Household Income** | | | | | |
| $150,000 and greater | 30 (38%) | 11 (37%) | 0.900 | Chi-square | |
| $100,000 to 149,999 | 15 (19%) | 5 (17%) | 0.780 | Chi-square | |
| $75,000 to 99,999 | 5 (6%) | 2 (7%) | 1.000 | Fisher's exact | |
| $50,000 to 74,999 | 6 (8%) | 3 (10%) | 0.704 | Fisher's exact | |
| $25,000 to 49,999 | 8 (10%) | 6 (20%) | 0.203 | Fisher's exact | |
| **Living Situation (multi-select)** | | | | | |
| Large city | 27 (34%) | 14 (47%) | 0.229 | Chi-square | |
| Suburb near large city | 29 (37%) | 7 (23%) | 0.185 | Chi-square | |
| Small city or town | 21 (27%) | 9 (30%) | 0.721 | Chi-square | |
| **Antibiotic Use (past year)** | | | | | |
| Yes | 33 (42%) | 7 (23%) | 0.074 | Chi-square | |
| **Diet Type** | | | | | |
| Omnivore | 63 (80%) | 27 (90%) | 0.266 | Fisher's exact | |
| Vegetarian (eats fish) | 8 (10%) | (7%) | 0.724 | Fisher's exact | |
| **Supplement Use (past month)** | | | | | |
| Probiotic supplement | 19 (24%) | 8 (27%) | 0.777 | Chi-square | |
| Fiber supplement | 13 (16%) | 5 (17%) | 1.000 | Fisher's exact | |
| **Bowel Movement Frequency (per day)** | | | | | |
| One | 49 (62%) | 17 (57%) | 0.609 | Chi-square | |
| Two | 16 (20%) | 9 (30%) | 0.280 | Chi-square | |
| Three | 4 (5%) | 4 (13%) | 0.212 | Fisher's exact | |
| **Stool type (Bristol Stool Scale)** | | | | | |
| Normal formed stool (Type 3-4) | 62 (79%) | 25 (83%) | 0.573 | | Chi-square |
| Loose/watery stool (Type 5-7) | 6 (8%) | 5 (17%) | 0.171 | | Fisher's exact |
| **GI Symptoms (past 30 days, multi-select)** | | | | | |
| Constipation | 30 (38%) | 15 (50%) | 0.255 | Fisher's exact | |
| Nausea | 12 (15%) | 4 (13%) | 1.000 | Fisher's exact | |
| **Diarrhea** | **27 (34%)** | **20 (67%)** | **0.002** | **Chi-square** | |
| Indigestion | 15 (19%) | 6 (20%) | 0.905 | Chi-square | |
| **Defecation discomfort frequency (past 30 days)** | | | | | |
| **0 times** | **46 (58%)** | **9 (30%)** | **0.008** | **Chi-square** | |
| 1–3 times | 21 (27%) | 12 (40%) | 0.173 | Chi-square | |
| 4–6 times | 7 (9%) | 4 (13%) | 0.491 | Fisher's exact | |
| 7–10 times | 3 (4%) | 2 (7%) | 0.614 | Fisher's exact | |
| More than 10 times | 2 (3%) | 2 (7%) | 0.303 | Fisher's exact | |

Data are presented as mean ± SD for continuous variables and n (%) for categorical variables. Categorical variables were compared using chi-square test or Fisher's exact test where expected cell counts were <5. Continuous variables were compared using independent samples t-test, with Mann-Whitney U (MWU) reported where distributional assumptions were violated. Race and living situation were multi-select items; percentages may not sum to 100%. Bold text indicates p<0.05. Abbreviations: SD, standard deviation; MWU, Mann-Whitney U test; GI, gastrointestinal.

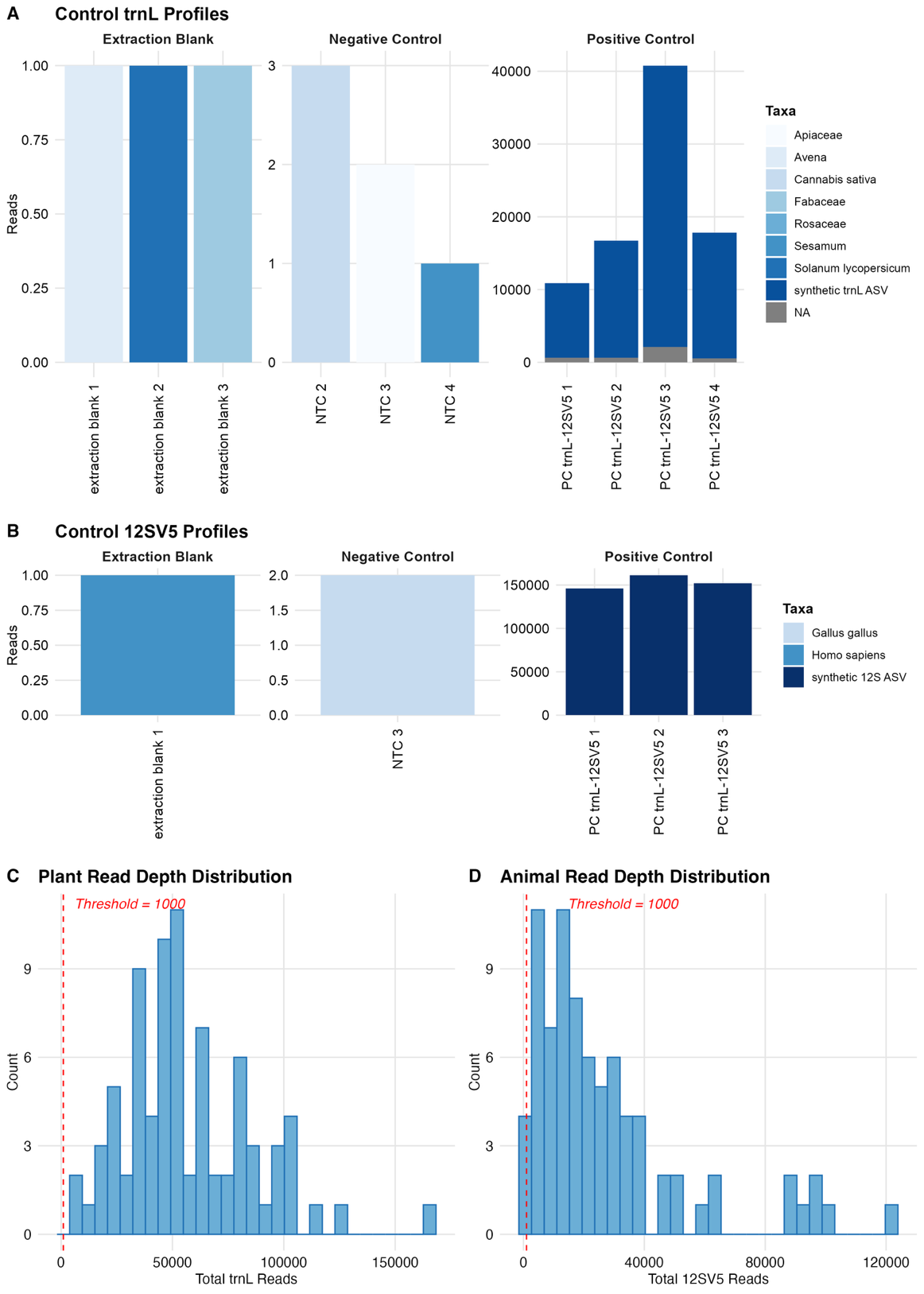

**Supplementary Figure 1 FoodSeq quality control metrics.** (**A**) trnL and (**B**) 12S V5 control profiles from a representative sequencing batch. Each batch included three extraction blanks, four no-template controls (NTCs), and three to four positive controls (PCs). Controls with zero reads are not displayed. Extraction blanks and NTCs with detectable reads showed minimal read counts, confirming low levels of reagent and environmental contamination. Positive controls contained the expected synthetic spike-in amplicon sequence variant (ASV) used as an internal standard. (**C**) Distribution of total trnL and (**D**) 12S V5 read depth across all participant stool samples. The dashed red line in each panel indicates the minimum read-depth threshold of 1,000 reads used to define sequencing success.

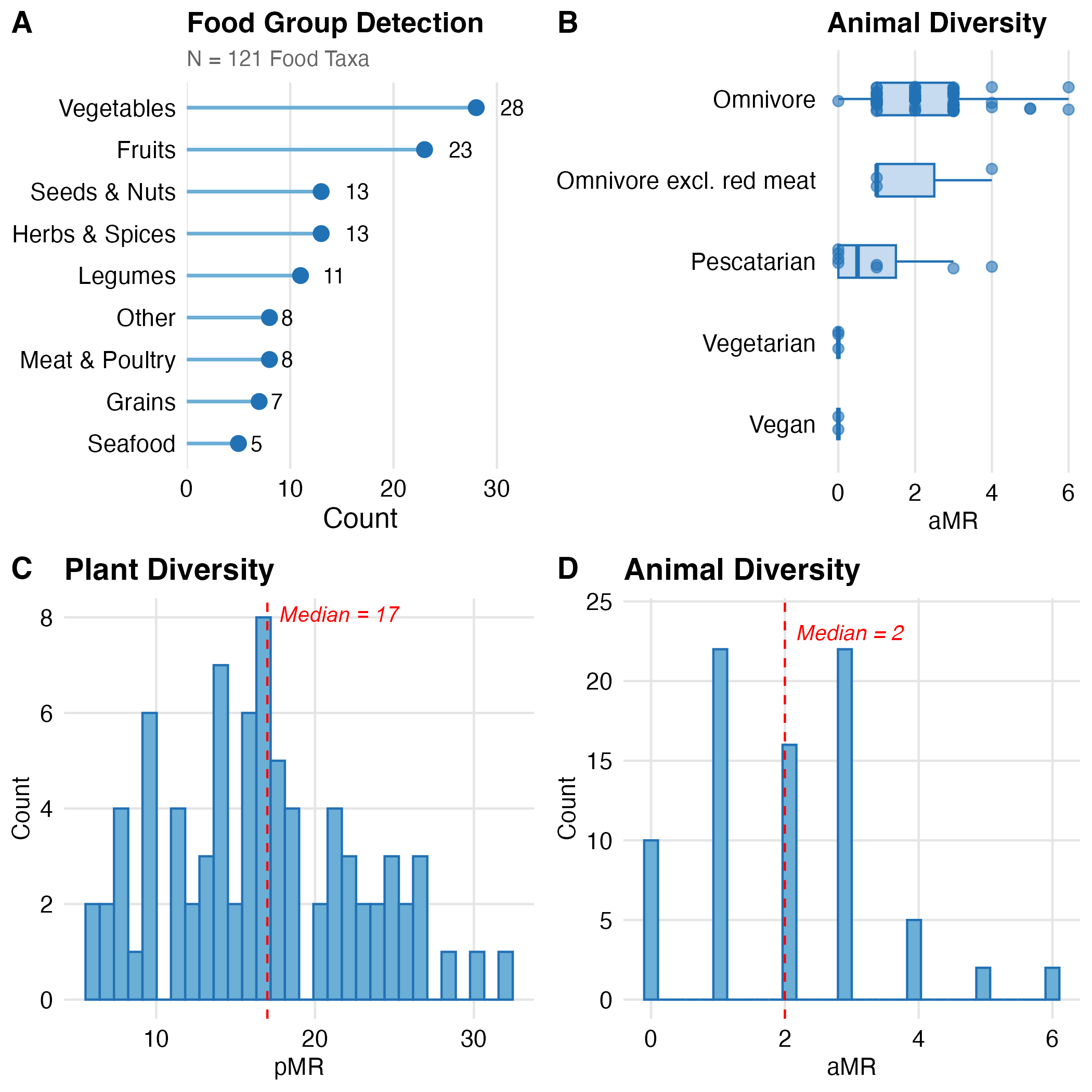

**Supplementary Figure 2 FoodSeq Dietary Diversity Metrics in the Everyone EATS cohort.** (**A**) Number of food taxa detected across nine food groups among participants with successful FoodSeq sequencing (N = 121 food taxa; N = 80 participants). Bars represent the total count of distinct taxa identified within each food group across the cohort. (**B**) Distribution of animal metabarcoding richness (aMR) by self-reported diet type. Box plots display the median, interquartile range, and range; individual data points are shown as jittered dots. (**C**) Distribution of plant metabarcoding richness (pMR) across all participants. pMR is defined as the count of distinct plant taxa detected per fecal sample using the chloroplast trnL-P6 marker. The dashed red line indicates the cohort median (pMR = 17). (**D**) Distribution of aMR across all participants. aMR is defined as the count of distinct vertebrate animal taxa detected per fecal sample using the mitochondrial 12SV5 marker. The dashed red line indicates the cohort median (aMR = 2).
